# Association of Parental Prenatal Mental Health on Offspring Neurodevelopmental Disorders: A Systematic Review and Meta-Analysis

**DOI:** 10.1101/2024.09.12.24313571

**Authors:** Adrianna P. Kępińska, Shelby Smout, Thalia K. Robakis, Lily E. Cohen, Ingrid Christina Gustavsson Mahjani, Alkistis Skalkidou, Veerle Bergink, Behrang Mahjani

**Affiliations:** Seaver Autism Center for Research and Treatment, Icahn School of Medicine at Mount Sinai, New York, NY, USA; Department of Psychiatry, Icahn School of Medicine at Mount Sinai, New York, NY, USA; Department of Genetics and Genomic Sciences, Icahn School of Medicine at Mount Sinai, New York, NY, USA; Department of Women’s and Children’s Health, Uppsala University, Uppsala, Sweden; Department of Psychiatry, Erasmus Medical Center, Rotterdam, The Netherlands; Department of Artificial Intelligence and Human Health, Icahn School of Medicine at Mount Sinai, New York, NY, USA; the Mindich Child Health and Development Institute, Icahn School of Medicine at Mount Sinai, New York, NY, USA; the Department of Medical Epidemiology and Biostatistics, Karolinska Institutet, Stockholm, Sweden; and the Department of Molecular Medicine and Surgery, Karolinska Institutet, Stockholm, Sweden

**Keywords:** anxiety disorders, obsessive-compulsive disorder, mood disorder, autism, neurodevelopmental disorders, attention-deficit/hyperactivity disorder, intellectual disability, maternal, paternal, prenatal

## Abstract

**Objective:** Parental prenatal mood and anxiety disorders (PMAD) are linked to child neurodevelopmental disorders (NDDs), but evaluations of the magnitude and mechanisms of this association are limited. This study estimates the strength of the association and whether it is impacted by genetic and environmental factors.

**Method:** A systematic search of PubMed, CENTRAL, PsycINFO, OVID, and Google Scholar was performed for articles published from January 1988 to January 2024. Of 2,170 articles screened, 64 met the inclusion criteria. Meta-analyses were conducted on 20 studies, and 44 were included in the narrative synthesis. We conducted random-effects meta-analyses, along with tests for heterogeneity (I²) and publication bias (Egger’s test). The review followed PRISMA and MOOSE guidelines.

**Results:** Maternal PMADs were associated with a significantly increased risk of ADHD (OR 1.91, 95% CI 1.45–2.52) and ASD (OR 1.57, 95% CI 1.37–1.81) in children. Paternal PMADs were also associated with the risk of NDDs, with combined odds for ASD and ADHD (OR 1.24, 95% CI 1.15–1.34). Several studies suggested that the link between parental PMADs and offspring NDDs might be impacted by both genetic and environmental factors, including the impact of ongoing parental depression on child behavior.

**Conclusions and Relevance:** Parental PMADs are significantly associated with an increased risk of NDDs in children. These associations may be influenced by both genetic predispositions and environmental factors. Understanding these pathways is important for informing interventions aimed at mitigating mental health risks in families and supporting child development.

## Introduction

As of 2021, 8.56% of children in the U.S. have a neurodevelopmental disorder (NDD), including autism spectrum disorder (ASD), intellectual disability (ID), and attention-deficit/hyperactivity disorder (ADHD). ^1,2^ Recognizing factors that predispose individuals to NDDs and enhancing early detection can lead to interventions that reduce the severity of NDD symptoms and improve the quality of life for neurodivergent individuals and their families.^3^ Furthermore, timely interventions can help prevent the progression of these conditions into other severe mental or medical complications in adulthood.

Recent research has demonstrated a significantly increased risk of NDDs in children born to mothers who experienced mood or anxiety disorders during pregnancy.^4,5^ This finding highlights the critical implications of maternal mental health on early neurodevelopmental outcomes and underscores the importance of integrating maternal health considerations into early pediatric care to support healthy developmental trajectories. Additionally, this body of research suggests a genetic link between ASD, ADHD, anxiety, and mood disorders, indicating that genes inherited from parents may predispose children to NDDs. It is also possible that genetic predispositions could interact with environmental factors such as maternal mental health to influence the development and severity of NDDs in children, thus necessitating a multifaceted approach in research and intervention strategies that consider both genetic and environmental contributions.

Despite extensive investigation into maternal mental health, such as pre and postnatal depression, significant gaps remain in our understanding, particularly the magnitude of the association and specific mechanisms through which prenatal mood and anxiety disorders (PMADs) affect neurodevelopmental outcomes. Additionally, while maternal influences have been extensively studied, the potential effects of paternal mental health remain largely unexplored. This oversight represents a critical gap in our knowledge, given the potential influence of paternal mental health and genetic contribution on developmental trajectories.

To address these gaps, we conducted a systematic review and meta-analysis of studies exploring the associations between both maternal and paternal PMADs—specifically prenatal depression, anxiety disorders, and obsessive-compulsive disorder (OCD)—and the risk of NDDs in offspring. This emphasis on the prenatal period is crucial, given its relatively limited coverage in existing literature compared to the broader perinatal and early postnatal periods. For NDDs, our focus was on three major conditions: ASD, ADHD, and ID, due to their shared genetic and phenotypic overlaps. For studies not suitable for meta-analysis, we performed narrative syntheses to provide a comprehensive overview of the existing literature. For the purposes of this study, we define “maternal” as relating to the parent who carried the pregnancy and “paternal” as relating to the other biological parent.

A crucial aspect of our study is analyzing whether the association between PMADs and NDDs in offspring primarily stems from shared genetic variation. We consider the possibility that this association may not be directly due to the environmental or behavioral influences of having a parent with PMADs but could predominantly result from genetic predispositions passed from parents to children. Understanding this potential genetic basis is critical for discerning whether the observed association reflects modifiable risk factors or primarily represents inherited genetic vulnerability.

## Methods

This systematic review and meta-analysis followed recommendations of the Preferred Reporting Items for Systematic Reviews and Meta-Analyses (PRISMA) (Supplement S1) and of Meta-analysis of Observational Studies in Epidemiology (MOOSE) guidelines.^6^ The study protocol is registered with PROSPERO (ID=CRD42022370757).

### Search strategy and selection criteria

Figure 1 outlines our article selection process, including identification, screening, and eligibility assessment. Two researchers completed searches (LEC and BM).

**Figure 1.**
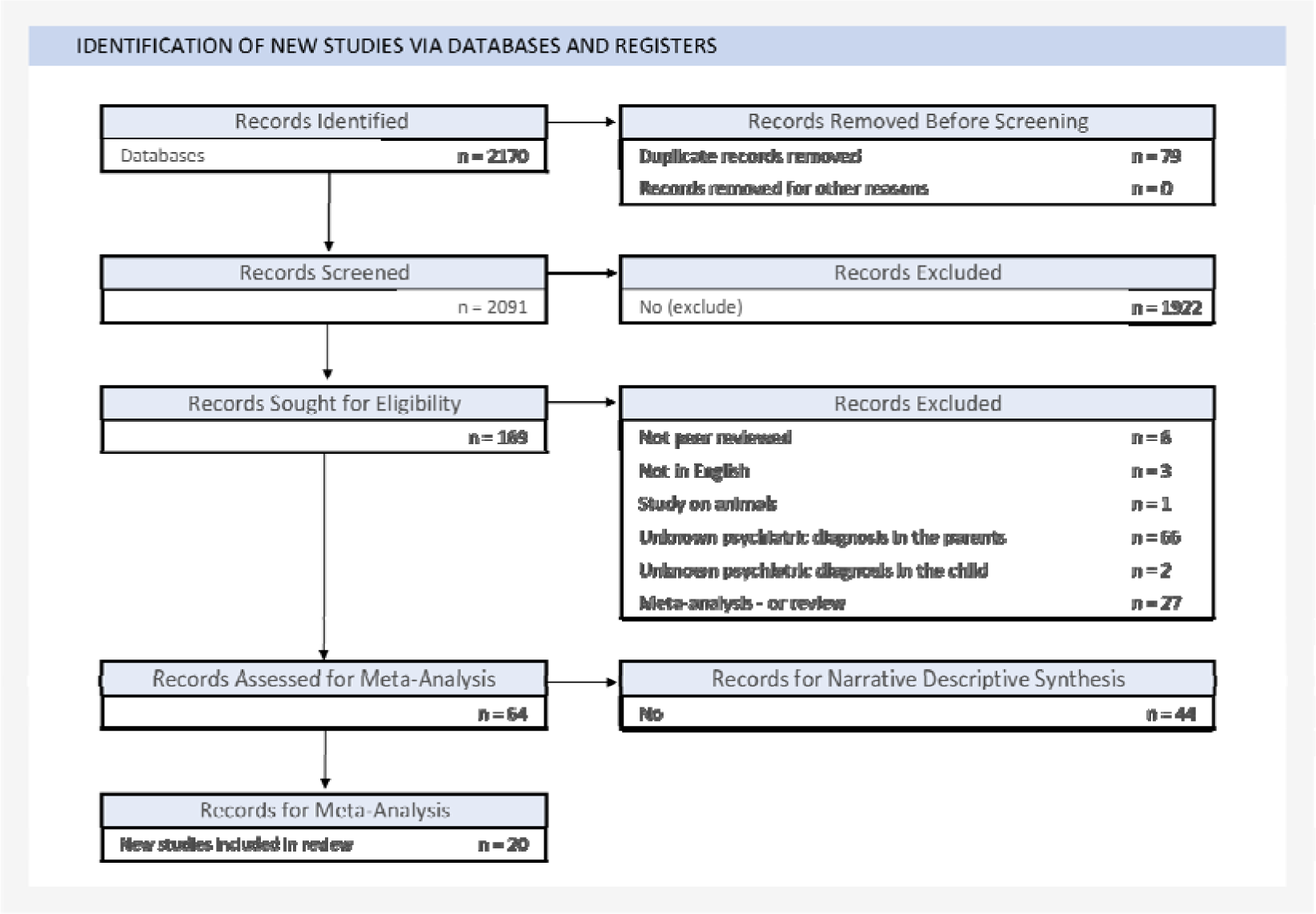
PRISMA flow diagram.

#### Inclusion Criteria

Studies on maternal/paternal PMADs and offspring NDDs were sought from PubMed, Cochrane CENTRAL, OVID, and Google Scholar, covering January 1, 1988, to January 16, 2024 (see Supplement S2 for search terms and details). We selected this study period to ensure comprehensive coverage of recent literature. We included cohort, case-control, and cross-sectional studies.

#### Exclusion Criteria

We excluded case studies, ecological/animal models, qualitative, and psychometric studies of PMADs and NDDs due to organic brain syndrome, substance use, or known physiological conditions. Since the goal of this study is to assess the relationship between parental PMADs and offspring NDDs, these studies were excluded to limit the number of confounding variables and pathophysiological mechanisms that could moderate this relationship. We also excluded grey literature, unpublished, not English-language research, and studies with fewer than 20 participants.

#### Screening

LEC and BM independently screened titles and abstracts to remove duplicates and irrelevant studies using DistillerSR software (Evidence Partners, Ottawa, Canada).

#### Eligibility

Full texts from screening and references from review articles were assessed independently by APK and LEC, with discrepancies resolved by discussion and random re-screening by BM. Articles were categorized based on whether they had relevant data for either the meta-analysis or narrative descriptive synthesis. The data required for inclusion in the meta-analysis is described below.

### Data extraction

Data extractions were independently conducted by three researchers (APK, LEC, and BM), with disagreements resolved through full-text review. Extracted data included: participant numbers with and without parents with PMADs and with and without NDDs; where available, reported measures of effect (odds ratios unadjusted for covariates, ORs; 95% confidence intervals, CI) and directions of effect; publication year; parental exposure to antidepressants; timing of parent exposure; parent age; parent exposure measures; offspring outcome (NDD diagnosis or symptoms); offspring age; sample size; number of offspring per sex/gender; setting; study/cohort/register name (where applicable); participant country of origin, ethnicity, race, or ancestry (as reported).

While our study primarily addressed the association of PMADs and NDDs, several studies examined the influence of prenatal antidepressant exposure. We only meta-analyzed or reviewed findings where antidepressants were ascertained as a treatment for PMADs and not other conditions, such as migraines and sleep disorders (Table S1).

### Statistical analysis

Our meta-analysis included parents diagnosed with mood or anxiety disorders during pregnancy or both prenatally and postnatally, excluding those diagnosed only postnatally. Where multiple studies used the same cohort, we selected the largest one to avoid sample overlap. We used a random-effects model for the meta-analyses to account for error variance within and between studies.^7,8^ We conducted the meta-analyses using R packages *meta* and *dmetar.* Effect sizes (OR) and variances were required for each study; for studies with counts of exposed and unexposed participants, parameters were calculated using the *meta metabin* function. When only unadjusted ORs and their confidence intervals (CIs) were available, the variance was derived from these CIs using the standard formula to convert CI width to standard error.

Using the R package *dmetar*, we assessed study heterogeneity with Cochran’s Q test, the I² statistic, and checked for outliers with funnel plots. Publication bias was addressed with the trim-and-fill method, which corrects for asymmetry in the funnel plot.^9^ We analyzed maternal and paternal findings separately and stratified analyses by offspring NDD for maternal studies only since the number of paternal PMAD studies was too low for stratification. A sensitivity analysis was conducted by meta-analyzing the data after excluding potential outlier studies.

Studies with information insufficient for meta-analysis were included in a narrative synthesis (see Supplement S3 for methods). Two researchers (APK and BM) independently assessed the risk of bias in all studies using the Newcastle-Ottawa scales and the Joanna Briggs Institute (JBI) Critical Appraisal Checklists (Supplement S4).^10–12^ Authors resolved any disagreements through discussion.

## Results

A total of 2,170 studies initially met the inclusion criteria. We selected 64 studies for the systematic review and 20 for meta-analysis (Figure 1).

Characteristics of meta-analyzed studies are detailed in Table 1. 27 studies reported participant race, country of origin, and ancestry or ethnicity, but none specified how these demographics were ascertained. The studies varied in their designs and implemented measurement tools (Table 1; additional details in Table S1).

**Table 1a.**
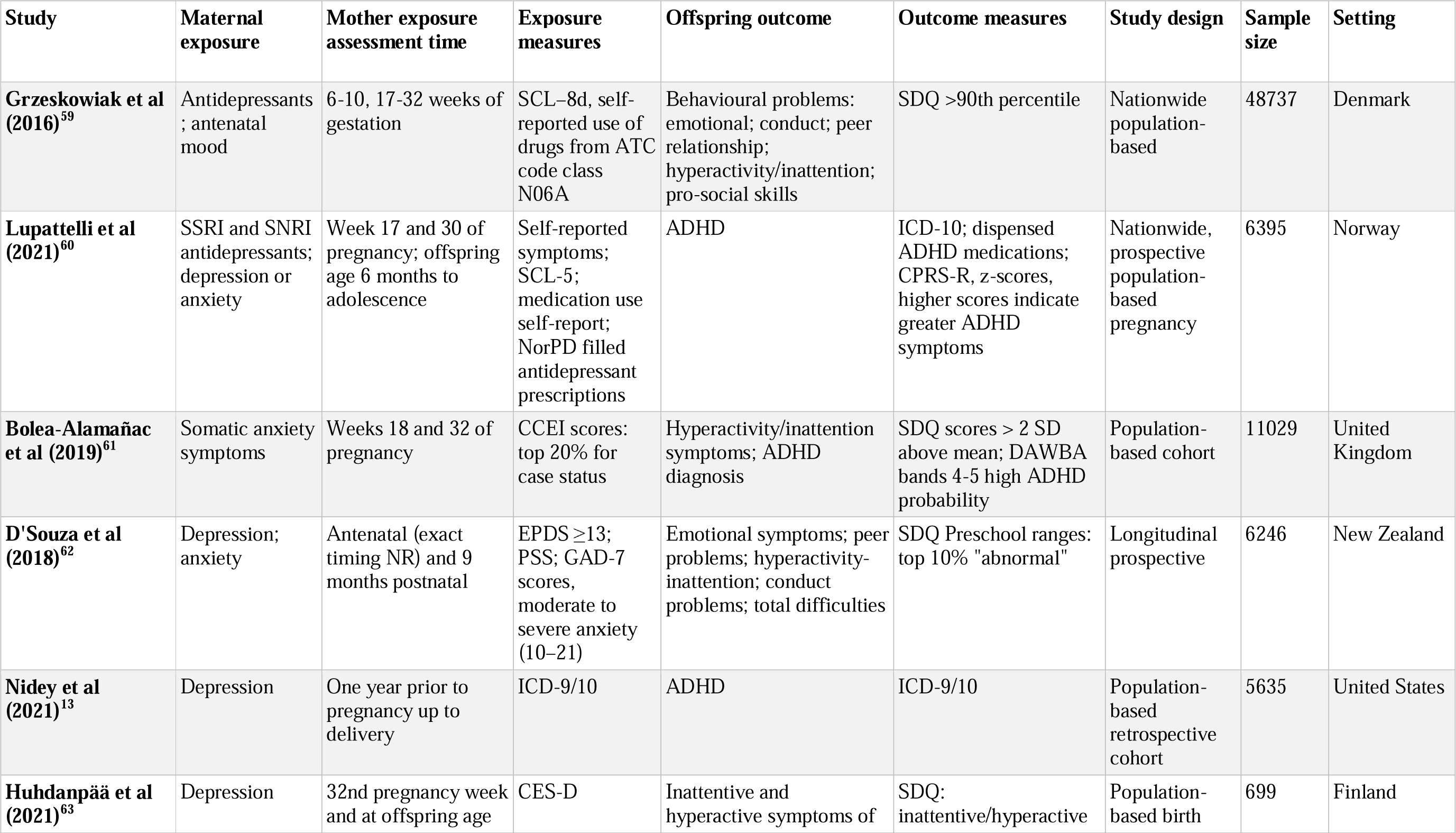
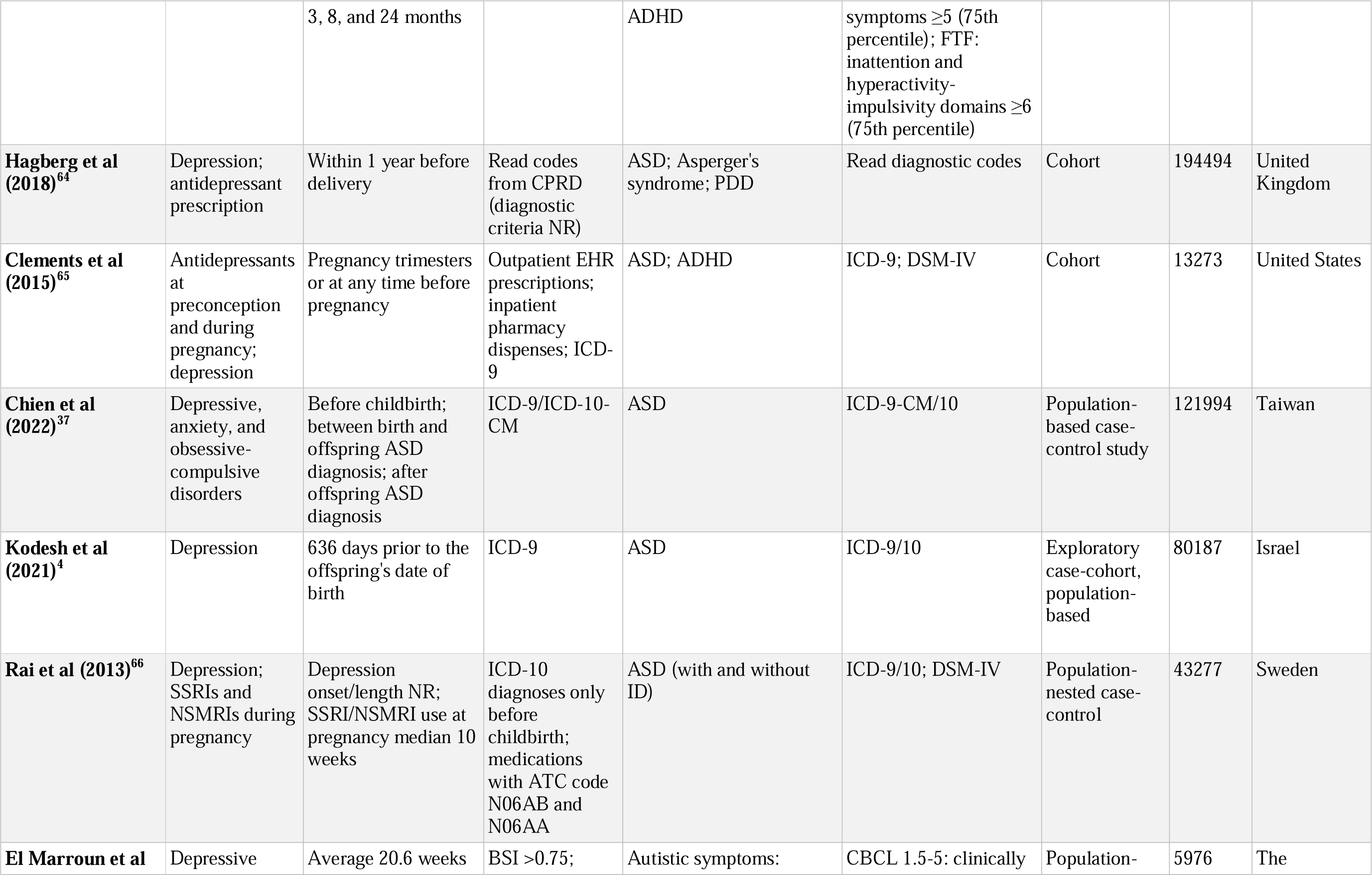
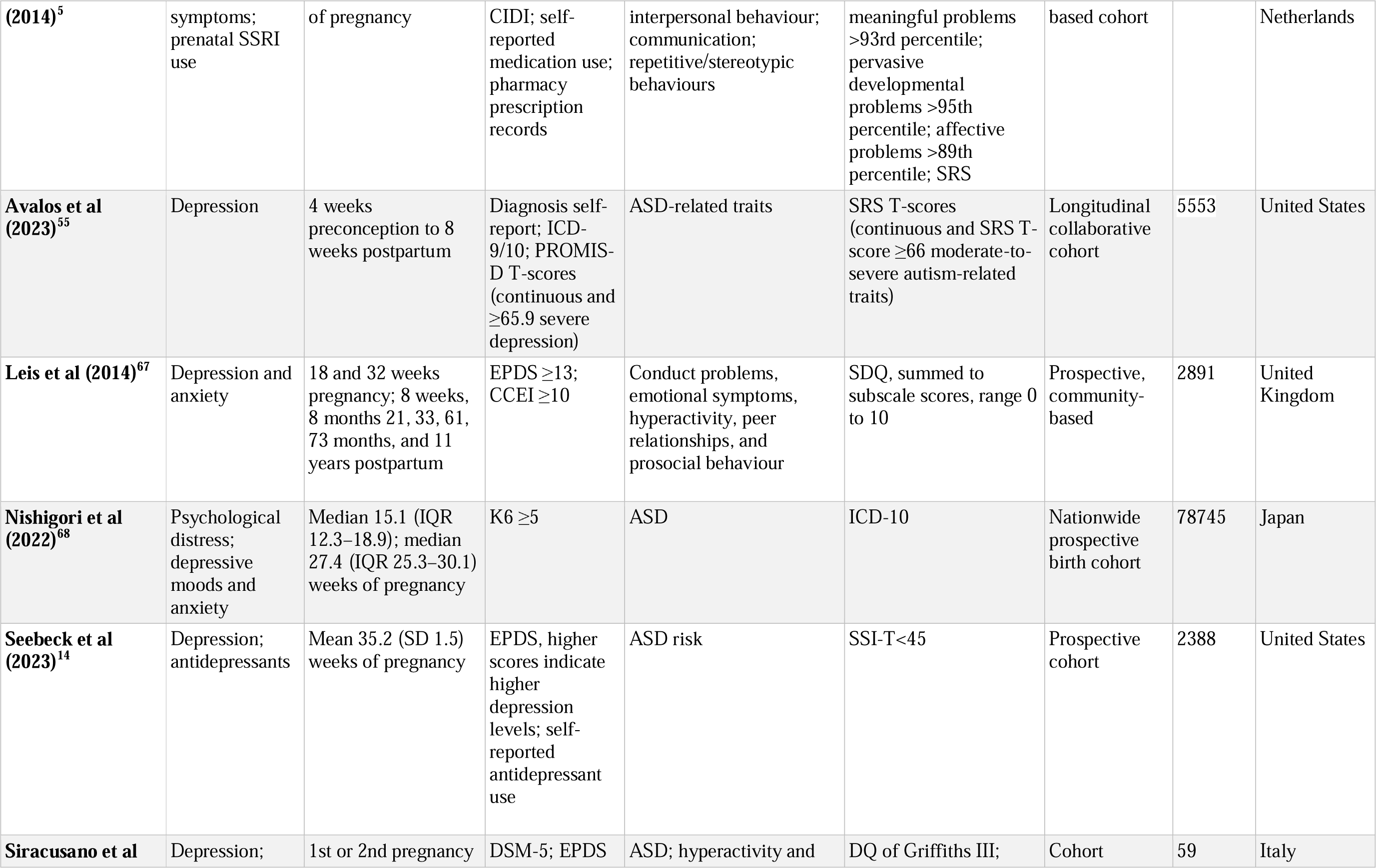
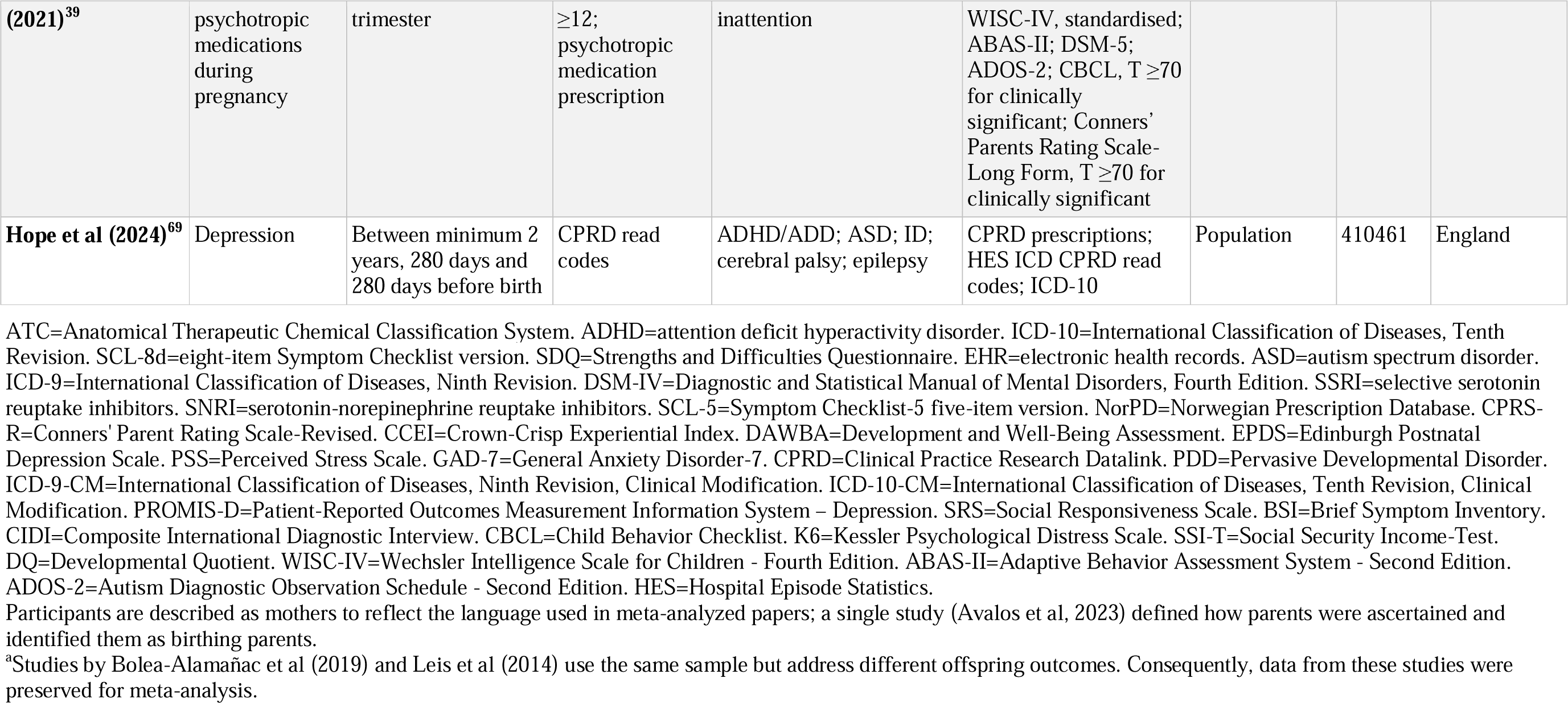
Characteristics of meta-analyzed studies on maternal prenatal mood and anxiety disorders and offspring neurodevelopmental disorders.

**Table 1b.**
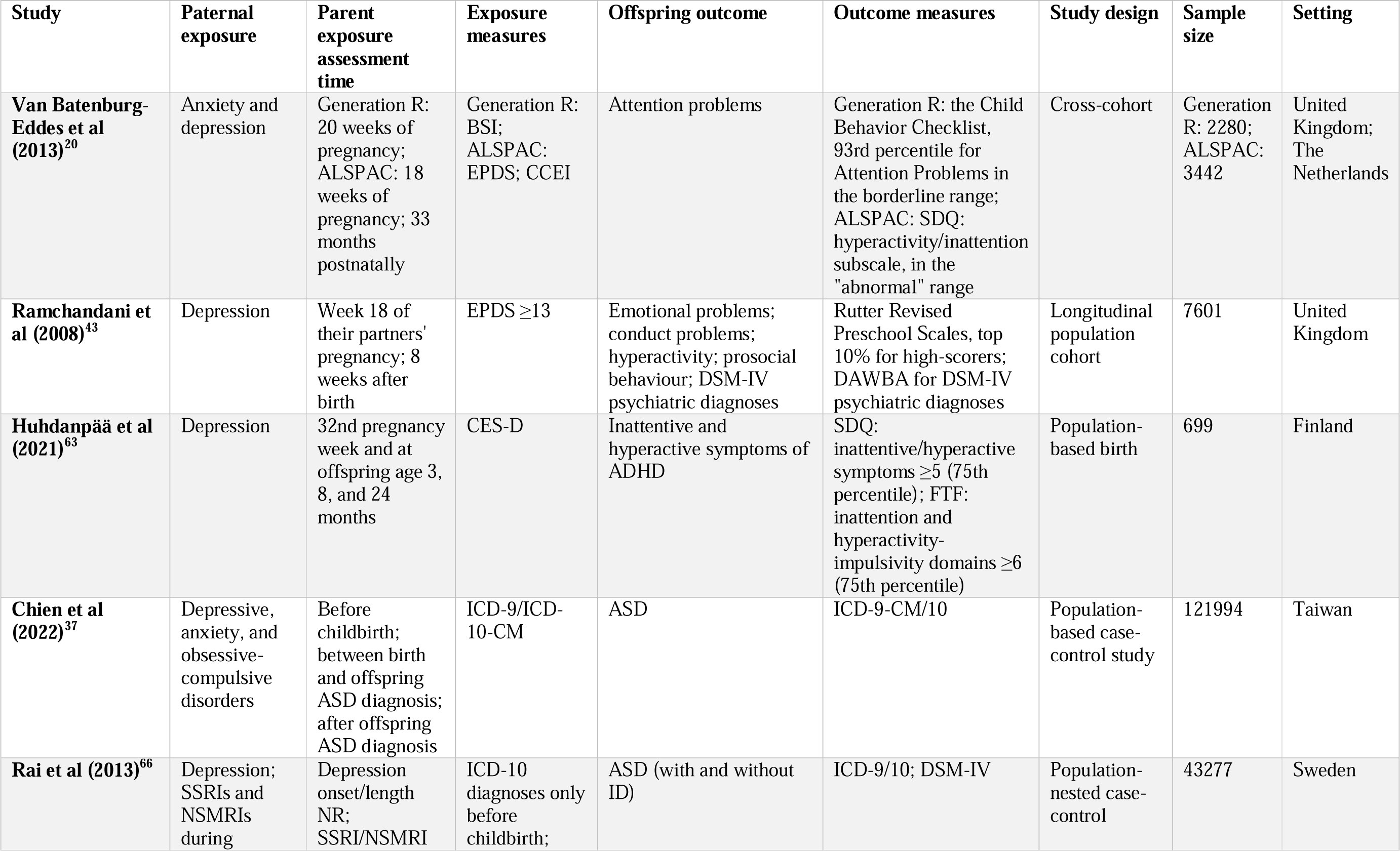
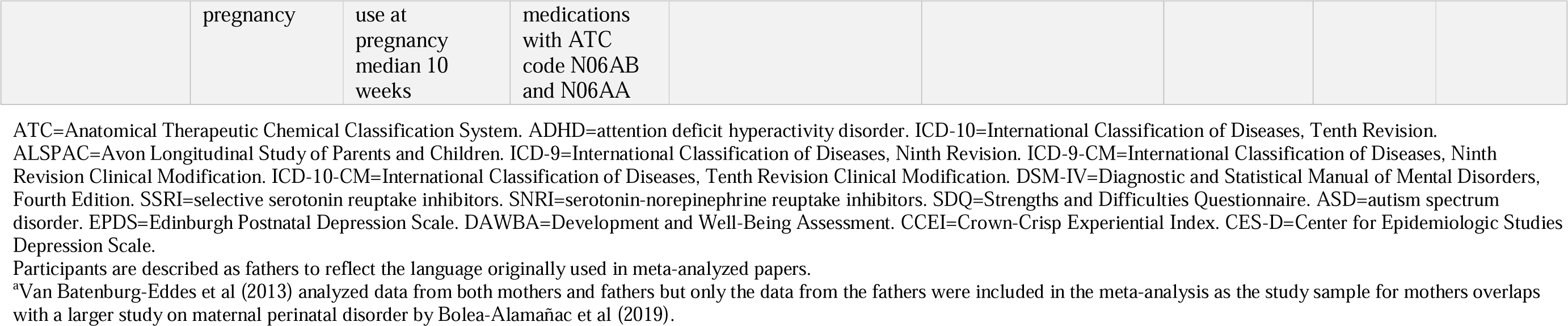
Characteristics of meta-analyzed studies on paternal prenatal mood and anxiety disorders and offspring neurodevelopmental disorders.

We additionally conducted a comprehensive narrative synthesis incorporating findings from the remaining 40 studies that were unsuitable for meta-analysis due to insufficient data for pooling. The findings from these studies are presented below.

### Maternal PMAD and the risk of offspring NDDs

Our meta-analysis revealed a significant association between maternal PMADs and offspring NDDs (OR 1.67, 95% CI 1.47–1.90, p<0.0001; Figure 2). Visual inspection of the funnel plot (Figure S1) identified two studies as possible outliers.^13,14^ Removing these studies reduced heterogeneity to low (Cochran’s Q=187.64, p<0.001 to Q=20.63, p=0.19; I^2^=90.4% to I^2^=22.5%) and OR to 1.59 (95% CI 1.50–1.68, p<0.0001; Figure S2).^15^

**Figure 2.**
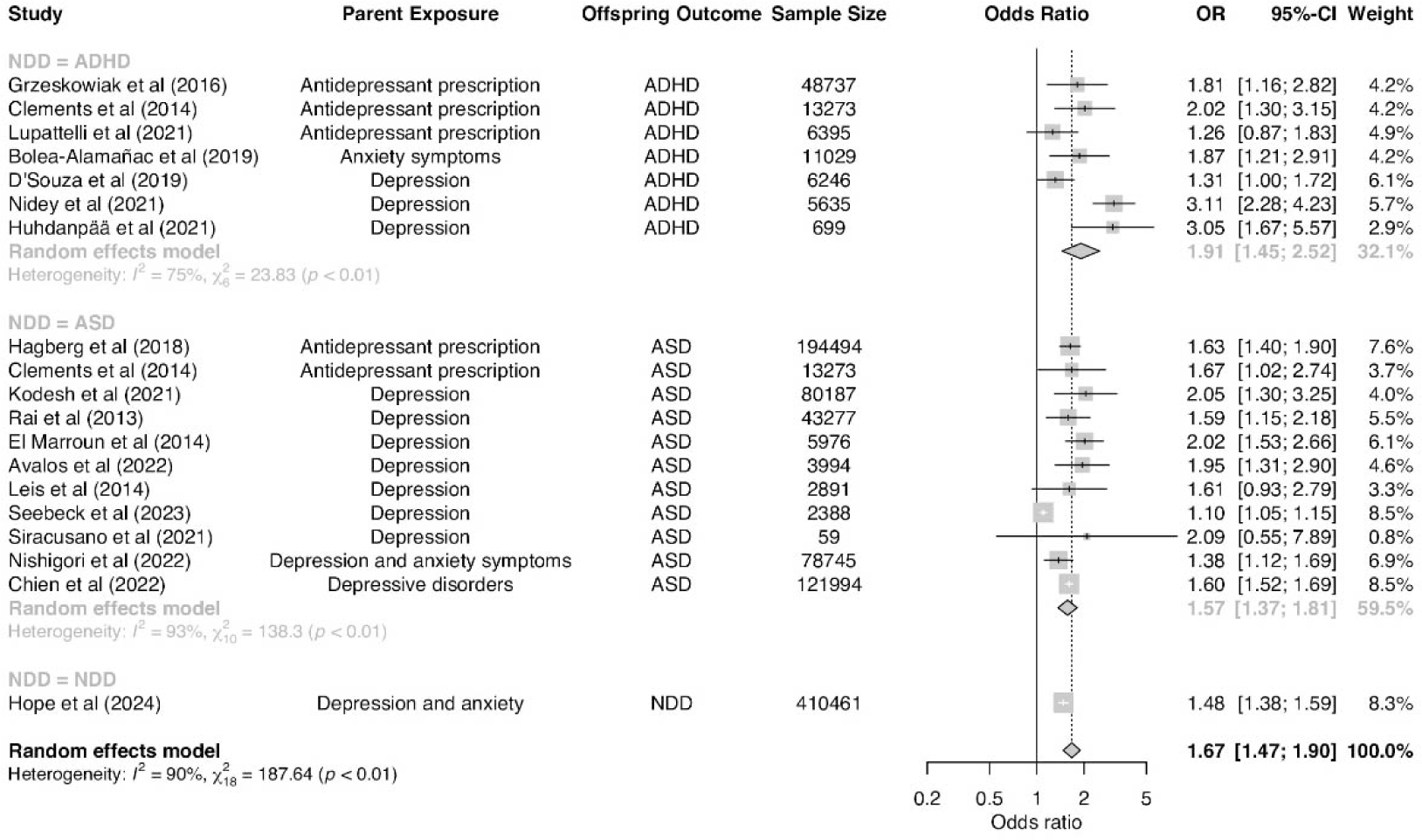
Forest plot of associations between maternal prenatal mood and anxiety disorders and offspring neurodevelopmental disorders. Square sizes reflect the weights attributed to each study. Diamonds denote the summary effect sizes for the random-effect models. OR=odds ratio. CI=confidence intervals NDD=neurodevelopmental disorders. ADHD=attention-deficit/hyperactivity disorder. ASD=autism spectrum disorder. ^a^Clements et al (2015) analyzed separate samples of individuals with diagnoses of autism spectrum disorder and attention-deficit/hyperactivity disorder. The authors provided results for three trimesters. The third trimester has been selected for this meta-analysis because it is the most conservative estimate reported by Clements et al (2015). ^b^Chien et al (2022) included the following disorders: major depressive disorder, persistent depressive disorder, and depressive disorder, unspecified. ^c^Hope et al (2024) analyzed a combined sample of individuals with any of the following disorders: autism/autism spectrum disorder, attention-deficit/hyperactivity disorder, intellectual disability, cerebral palsy, and epilepsy.

The Egger’s test indicated asymmetry in the funnel plot (p=0.03; Figure S1), which suggests publication bias. In support of this result, the trim-and-fill method estimated at least nine potentially missing studies (Figure S3).

### Effects of maternal PMADs on offspring ADHD

Our meta-analysis revealed a significant association between maternal PMADs and an increased risk of offspring ADHD (OR 1.91, 95% CI 1.45–2.52; Figure 2). Initially, the ADHD model exhibited high heterogeneity (Cochran’s Q=23.83, p<0.001; I^2^=74.8%). After the exclusion of a single outlier study, heterogeneity decreased (Cochran’s Q=9.90, p=0.08; I^2^=49.5%), and OR changed to 1.70 (95% CI 1.34–2.14; Figure S1).^15^

Consistent with the meta-analysis, studies in the narrative synthesis reported associations between maternal PMADs and offspring ADHD or elevated ADHD symptoms.^16^ However, the literature presented mixed results for the mechanism underlying this finding. LM Chen and colleagues employed polygenic risk scores (PRS) for ADHD and demonstrated that a significant portion of the association between maternal prenatal depression and ADHD in offspring could be attributed to shared genetic risk factors between these disorders.^17^ In contrast, Eilertsen et al. demonstrated a persistent positive association between maternal prenatal depression symptoms and the risk of ADHD in offspring, even after controlling for shared genetic factors.^18^ Adding to this complexity, Olstad et al. investigated epigenetic factors and found no association between cord blood DNA methylation, maternal prenatal depression, and offspring ADHD symptoms.^19^Similar to maternal depression, maternal anxiety has been associated with an increased risk of offspring ADHD, but studies remain inconclusive on the critical exposure period. One study found only maternal anxiety at 12-22 weeks gestation, but not later, predicted offspring ADHD symptoms, while another highlighted that interaction between maternal anxiety at 32 weeks and offspring genes influences ADHD symptom severity.^20,21^ This discrepancy may be due to differing study designs, particularly the inclusion of genetic interactions in the latter study.^21^While individual time points are informative, trajectory studies provide a more comprehensive picture of the relationship between maternal PMADs and offspring ADHD. Trajectory studies of maternal depression suggested that either persistently high or increasing levels of maternal depression during pregnancy were associated with increased offspring ADHD symptoms.^22–26^ Similarly, consistently low or decreasing levels of maternal depressive symptoms have been associated with fewer offspring ADHD symptoms, with reductions in maternal symptoms corresponding to decreases in offspring symptoms over time.^22–24^ This pattern indicates that both the timing and the severity of maternal depression are important factors, although the precise mechanisms are not yet fully understood.

Finally, research comparing the association of anxiety and depression found that increased maternal anxiety correlated with higher child hyperactivity, while changes in maternal depression were related to all ADHD symptoms except inattention.^27^ However, these patterns have not been consistently reported across studies, possibly due to variations in sample sizes and study designs.^28–33^

### Effects of maternal PMADs on offspring ASD and ID

Our meta-analysis revealed an association between maternal PMADs and an increased risk of offspring ASD (OR 1.57, 95% CI 1.37–1.81). Studies initially presented with high heterogeneity (Cochran’s Q=138.30, p<0.001; I^2^=92.8%), but upon removal of one study, heterogeneity was low (I^2^=0%; Cochran’s Q=6.96 p=0.64). OR increased to 1.61 (95% CI 1.54–1.69; Figure S2).^14^

Narrative synthesis largely corroborated the finding that maternal PMADs increase the risk of ASD and autistic-like traits in offspring.^34^ One study found that children of mothers with PMADs were significantly more likely than children of unaffected mothers to exhibit mild language and motor delays rather than severe developmental delays, with the risk intensifying when mothers had both depression and anxiety pre-delivery. However, this finding should be interpreted with caution, as anxiety was based on a self-reported questionnaire and the study included controls with common comorbidities, such as bipolar disorder.^34^

Further supporting these associations, an analysis of cord blood from mothers with prenatal depression or post-traumatic stress disorder found increased expression of genes associated with ASD.^35^ These genes were linked to reduced cognitive performance in their two-year-old infants. These findings suggest that maternal PMADs may influence infant brain development through altered neurodevelopmental pathways beyond directly inherited genetic risk.

Studies on OCD were rare. A single study addressed maternal OCD and reported a significant association between maternal OCD within four years before delivery and offspring ASD (adjusted OR = 3.42, 95% CI = 1.77–6.63).^36,37^ Although this study included the entire Taiwanese population, it only involved 12 OCD cases and 36 controls, highlighting the need for additional research with larger sample sizes to support these findings.

Several observational studies initially suggested an association between ASD and antenatal antidepressant exposure, but these studies lacked adequate controls. ^14,38,39^ Conversely, studies that included groups of siblings discordant for antidepressant exposure have established that the association is not related to any direct pharmacological effect but, rather, the shared genetic and environmental risk factors related to the indication for prescribing antidepressants (i.e., maternal depression and related symptoms).^40–42^ Furthermore, one study suggested potential parental rater bias in evaluating the severity of child developmental problems among families with prenatal selective serotonin reuptake inhibitor (SSRI) exposure.^5^ Findings indicated an association between SSRI exposure and autism symptoms in children when both parents provided reports on child development. However, when only fathers reported, there was no significant association between maternal depression during pregnancy and child developmental problems. This finding indicates that parental reports might not always be reliable, potentially leading to biased estimates of children’s behavioral issues.

### Paternal PMAD and the risk of offspring NDDs

Our meta-analysis identified a significant association between paternal PMADs and offspring NDDs (OR 1.24, 95% CI 1.15–1.34, p<0.001; Figure 3). Given the few available studies, it was not possible to conduct separate meta-analyses on the effects of paternal PMADs on ADHD and ASD. Studies showed low heterogeneity (Cochran’s Q=1.36, p=0.93; I^2^=0%) with no outliers.

**Figure 3.**
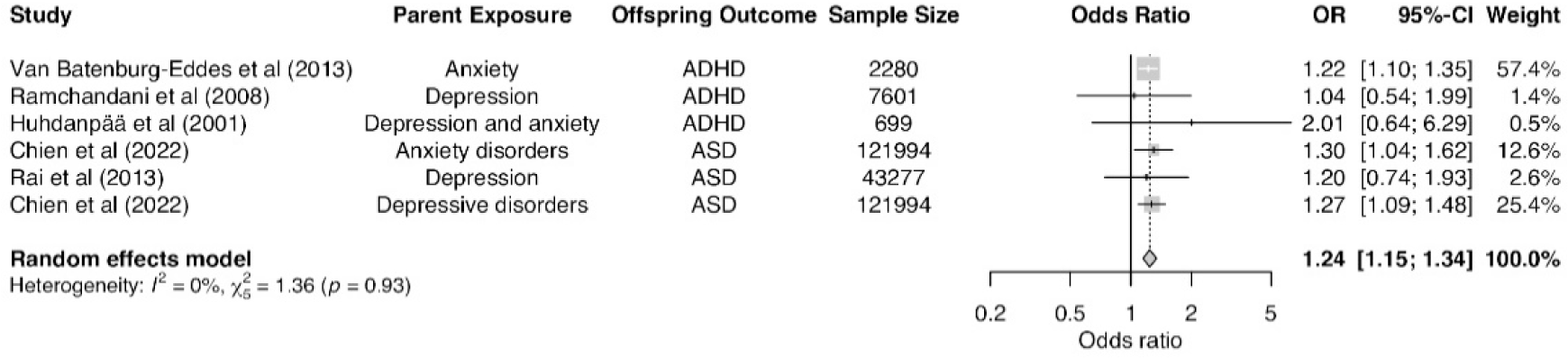
Forest plot for the association between paternal prenatal mood and anxiety disorders and the risk of neurodevelopmental disorders (NDDs) in offspring. Square sizes reflect the weights attributed to each study. The diamond denotes the summary effect size for the random-effect models. CI=confidence intervals. ASD=autism spectrum disorder. ADHD=attention-deficit/hyperactivity disorder. ^a^Chien et al (2022) included the following disorders: major depressive disorder, persistent depressive disorder, depressive disorder, unspecified, generalized anxiety disorder, panic disorder, agoraphobia, social anxiety disorder, and specific phobia disorder.

**Figure 4.**
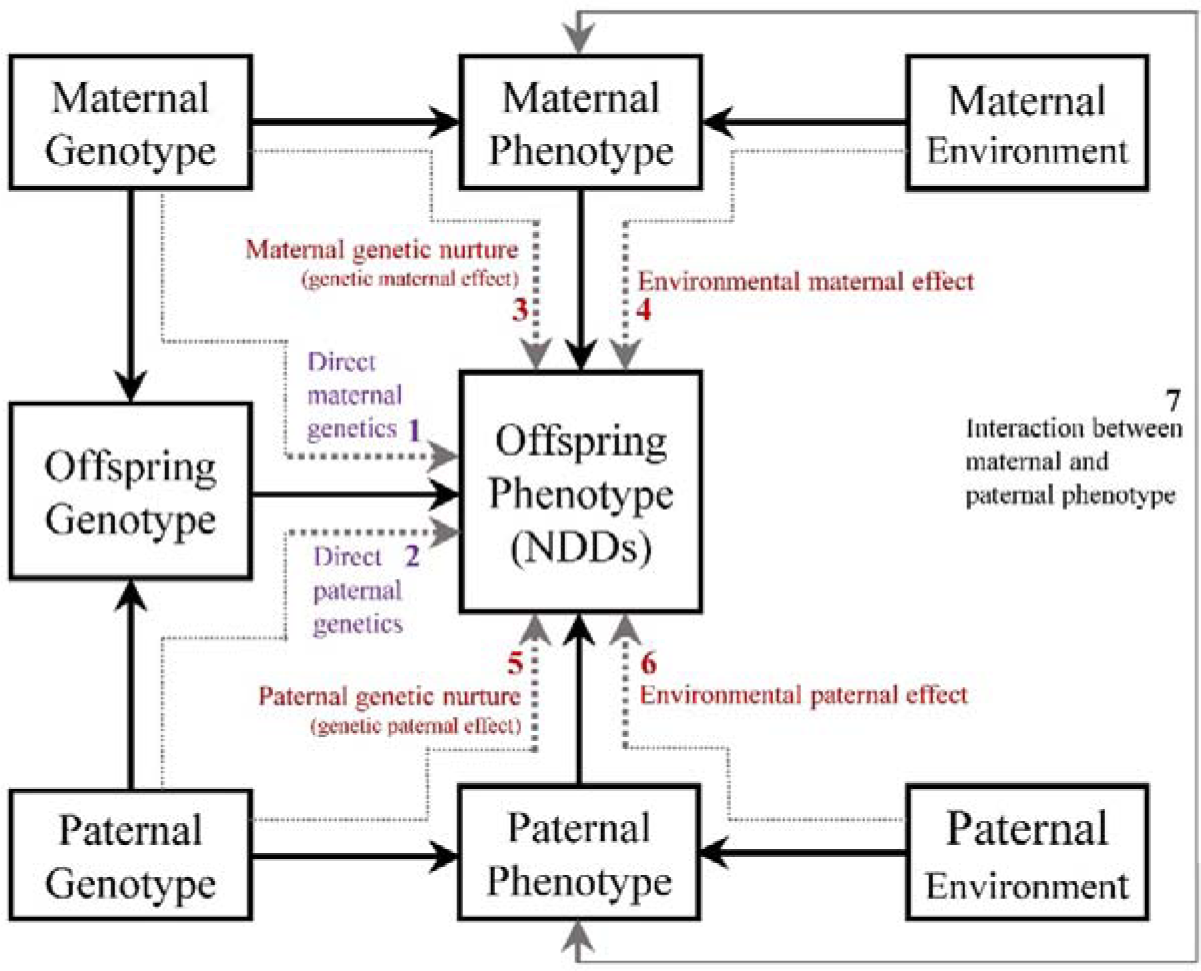
Maternal and paternal effect on offspring phenotype (a neurodevelopmental disorder). It includes three main pathways: the maternal/paternal genetic nurture effect, showing transmission from maternal/paternal genotype to their respective phenotypes and then to the offspring’s phenotype (dashed arrows 3 & 5); the environmental maternal/paternal effect, indicating the influence of the maternal or paternal environment on their phenotype and consequently on the offspring’s phenotype (dashed arrows 4 & 6); and the direct maternal/paternal genetic effect, which traces direct transmission from maternal/paternal genotype to the offspring’s genotype and then to their phenotype (dashed arrows 1 & 2). This figure omits child environment effects or gene-environment interactions.

The funnel plot (Figure S4) was asymmetrical, suggesting potential publication bias, but Egger’s test was non-significant (p=0.58). However, the number of studies may be too small to test for small study effects. The trim-and-fill method (Figure S5) indicated at least one missing study, suggesting unreported results that could affect the overall effect estimate.

### Effects of paternal PMAD on ADHD in offspring

Chen et al. showed that paternal prenatal depression increases offspring ADHD risk, particularly in cases of chronic depression or when both parents are affected.^17^ In contrast, Ramchandani et al. found no significant association between paternal depression during pregnancy and offspring ADHD diagnoses.^43^ Other studies also reported limited evidence for an association between prenatal paternal depression, anxiety, and risk of offspring ADHD.^21,25,31^

Adding to this complexity, a single study found that, after controlling for shared genetics, prenatal maternal depression was associated with a slightly elevated offspring ADHD risk, while prenatal paternal depression showed a minor association with lowered offspring ADHD risk.^18^ Nonetheless, researchers cautioned against drawing definitive conclusions from this association, noting the lack of theoretical models that explain the effect of paternal prenatal depression. In light of their findings, they also questioned the validity of selecting parents with depression as negative controls for one another in future studies.

### Effects of paternal PMADs on offspring ASD

Chen et al. found that offspring ASD risk was elevated when either parent was affected, with a slightly greater risk for paternal than maternal prenatal depression. Additionally, a separate study based on Taiwanese registers found no significant associations between paternal prenatal OCD and offspring ASD, likely due to small sample sizes.^36,37^

### Sex or gender differences

Few studies addressed interactions between offspring sex or gender, NDDs, and maternal and/or paternal PMADs. Of these few, none qualified for meta-analysis. The influence of prenatal maternal and paternal PMADs on NDDs appears to differ by sex, though the existing research is limited and yielded mixed results. The studies did not distinguish between sex and gender.

Loomans et al. observed that maternal prenatal anxiety significantly correlated with increased hyperactivity and inattention problems in boys but not in girls, suggesting potential sex-specific vulnerabilities to prenatal anxiety.^44^ In contrast, Huhdanpää et al. found a similar pattern, with maternal prenatal depressive symptoms linked to increased inattentiveness and hyperactivity among boys at age five.^25^ These findings imply that boys might be particularly susceptible to the neurodevelopmental influence of prenatal maternal affective symptoms.

Conversely, Bendiksen et al. reported that the higher prevalence of ADHD symptoms, particularly the hyperactive/impulsive subtype in boys, does not reflect a differential contribution of maternal distress between the sexes.^30^ This suggests that while boys generally display more ADHD symptoms, the effects of maternal distress during pregnancy affect both sexes similarly. Supporting this lack of sex-specific associations, Chen et al. found no sex differences in the association between prenatal depression and child mental health outcomes.^45^ They suggested that earlier findings indicating that prenatal maternal stress predicts sex-specific child outcomes may vary depending on the particular behavior being examined.

Studies found no sex/gender differences following exposure to maternal prenatal depression and autism-related traits or behavioral problems in offspring.^45^ No studies specifically addressed paternal PMADs.

### Risk of bias

According to assessments conducted with the Newcastle-Ottawa scales, the majority of studies on parental PMADs were at moderate risk of bias (36 studies, 59%), and five studies (8%) were at low risk. Among studies assessed with JBI checklists, two out of three studies reported key information and used correct statistical analyses (Table S2).

## Discussion

This systematic review and meta-analysis examined the relationship between PMADs in parents and the risk of NDDs in their offspring. We found a stronger association for maternal PMADs (OR 1.67, 95% CI 1.47–1.90) than paternal PMADs (OR 1.24, 95% CI 1.15–1.34).

Although not explored in this study, prior research suggests that this larger association with maternal PMADs might be attributed to more direct biological and environmental effects during pregnancy.^46,47^ Although our primary focus was on prenatal disorders, the observed associations might also reflect the influence of postnatal maternal mental health, as many women experience mood and anxiety disorders beyond childbirth.^48,49^

In the meta-analysis, we observed moderate heterogeneity after excluding two studies.^13,14^ Potential sources of heterogeneity include variation in sample sizes, diverse PMAD and NDD measurement tools, diverse timeframes between conception and delivery, and few studies accounting for prenatal treatment initiation.^50^

Our meta-analysis revealed a strong association between maternal depression and anxiety and an increased risk of ADHD in offspring. The narrative synthesis underscored that specific timing or trajectory of maternal disorder might have varying effects on offspring ADHD. Notably, maternal anxiety during early pregnancy (12-22 weeks) significantly predicted ADHD symptoms.^20^ This finding indicates the potential influence of factors beyond shared genetics, as a purely genetic link would likely yield consistent effects irrespective of the timing of maternal anxiety. Additionally, a single study highlighted a positive association between maternal prenatal depression symptoms and offspring ADHD risk after adjusting for genetic factors.^18^ Together, these findings suggest that prenatal environment may mediate the association between PMADs and offspring ADHD. The meta-analysis also revealed a strong association between maternal depression and anxiety and an increased risk of offspring ASD.^17,51,52^

Studies regarding prenatal OCD were limited and found an association between prenatal maternal OCD and offspring ASD.^36^ Studies of offspring ID were also scarce and either analyzed combined ID and NDDs, or ID-related symptoms. These studies reported that offspring of mothers with PMADs were at greater risk of mild language and motor delay or gross motor deficits.^53,54^

Finally, current research addressing the association of parental PMADs on sex/gender differences in offspring NDDs is inconsistent. Some studies suggest a tentative association with ADHD symptoms in boys, but not girls, and no significant gender/sex associations with ASD or autism-related traits.^25,30,44,55^ These differences may stem from gender-or sex-specific differences in how parental mental health influences child outcomes, methodological differences between studies, or cultural patterns that influence diagnosis. For instance, the ADHD symptom of hyperactivity may be more disruptive than inattention in certain socioeconomic and healthcare environments, leading to higher diagnostic rates in boys.

Alternatively, diagnostic rates may be affected by gender role expectations or stereotypes of neurodivergent presentations in individuals with different gender identities. Another explanation may be that more recent studies used larger samples, including testing multiple cohorts.^16,55^ Patterns of results may differ between diverse populations.

Multiple pathways are likely involved in the association between maternal PMADs and the risk of NDDs in offspring. The first pathway (Figure 2, arrow 1) involves direct transmission of genes from the mother to the offspring. The second and third pathways (arrows 3 & 4), are known as *maternal effect*.^56^ Maternal effect refers to the impact of the mother’s phenotype on the offspring’s phenotype, above and beyond the transmission of maternal genes: via environmental pathways instead of directly transmitted genetic risk. Maternal effect can arise from maternal genotype (maternal genetic effect, also called maternal genetic nurture, arrow 3), and/or maternal environment (environmental maternal effect, arrow 4). Paternal PMADs are also associated with the risk of offspring NDD via three analogous pathways (arrows 2, 5, 6).

Maternal/paternal effects have been defined as causal impacts of the maternal/paternal genotype or phenotype on the offspring phenotype.^56^ However, statistical models used for estimating parental effects may not always provide causal estimates due to several factors: (1) *Maternal/paternal effects could depend on offspring genotypes,* such as an additive-by-additive interaction between the maternal/paternal and the offspring genotype.^56^ For instance, an interaction between maternal anxiety and the offspring gene may predict the severity of ADHD symptoms, suggesting an additive-by-additive interaction between the maternal and offspring genotypes.^21^ (2) *Lack of blinding in observational family-based studies* may lead to diagnostic bias, as relatives of probands with psychiatric disorders may be more likely to be diagnosed, due to family history being part of the diagnostic process. This could result in overestimated genetic influence in family-based studies; (3) *Assortative mating* (non-random mating) can influence population prevalence estimates and estimates of direct genetic effect; (4) *Selection bias* due to differential access to healthcare, where individuals with milder disorder forms or who do not seek or cannot access clinical services may be excluded from research; (5) *Gene-environment interactions can complicate causal inference* (they were omitted in Figure 1). Several studies reviewed here suggested such mechanisms, e.g., for offspring ADHD, depression or anxiety interacting with smoking during pregnancy, prenatal infection with prenatal anxiety, and short breastfeeding in mothers with prenatal depressive mood or anhedonia;^27,30,51,57^ (6) *Measurement errors* can introduce uncertainties and biases into the analysis; (7) *Failure to achieve exchangeability between exposed and unexposed groups* may lead to confounding, undermining the validity of causal conclusions; and (8) *Retrograde effects*, where the offspring phenotype may influence parental phenotype. Examples include fetomaternal immune incompatibility, such as Rh factor incompatibility, which could lead to maternal depressive symptoms and later cognitive effects in the offspring.

### Strengths and limitations

The primary strength of our study is its pre-registered analysis of multiple studies from diverse regions, including Europe, North America, Asia, and Australia. However, the study also has several limitations. Due to limited available findings, the meta-analysis of paternal PMADs may have less statistical power than the findings on mothers. Furthermore, none of the studies provided explicit definitions of mothers and fathers, making it difficult to determine generalizability to transgender or non-binary parents, co-parents, or non-biological partners. Moreover, several studies come from the same populations, predominantly representing Western sites. Although we selected the largest sample sizes from studies with the same cohorts for meta-analyses, the narrative synthesis included findings from different studies that used the same cohorts. This approach may introduce unique limitations for each cohort. For instance, the Norwegian cohort MoBa has a low response rate (43.5%) and may suffer from self-selection of healthier participants into the cohort.^58^ Additionally, most studies do not report detailed breakdowns of ethnicity, race, or ancestry, limiting our ability to assess how globally representative the samples are.

### Conclusion

Our comprehensive meta-analysis, reinforced by narrative synthesis, supports the association between maternal and paternal PMADs and an increased likelihood of NDDs in offspring. This association may not be entirely attributable to set genetic predispositions passed from parents to children; rather, it could be partially moderated by the environmental or behavioral impacts of having a parent with PMADs. To reduce the severity of neurodevelopmental disorders (NDDs) and related symptoms, intervention strategies should consider both genetic and environmental factors. These strategies could involve genetic counseling to identify families who would benefit from early mental health screenings. This approach enables the modification of environmental influences through timely mental health support and effective stress management. Recognizing the importance of both genetic and environmental factors is crucial for developing successful treatment strategies for NDDs.

## Contributions

*Concept and design:* Robakis, Bergink, Mahjani

*Acquisition, analysis, or interpretation of data:* Kępińska, Robakis, Bergink, Mahjani

*Drafting of the manuscript:* Kępińska, Mahjani

*Critical revision of the manuscript for important intellectual content:* All authors

*Statistical analysis:* Kępińska, Mahjani

*Obtained funding:* Kępińska, Robakis, Bergink, Mahjani

*Supervision:* Bergink, Mahjani

## Data Availability

All data produced in the present work are contained in the manuscript

## Acknowledgments

This project was funded by the Beatrice and Samuel A. Seaver Foundation (Kępińska, Mahjani), Brain & Behavior Research Foundation (Mahjani), the National Institute of Mental Health (NIMH) R01HD111117 (Robakis), R21MH131933 (Kępińska, Robakis, Bergink, Mahjani)

## References

1. Boyle CA, Boulet S, Schieve LA, et al. Trends in the prevalence of developmental disabilities in US children, 1997-2008. Pediatrics. 2011;127(6):1034-1042.

2. Zablotsky B, Ng A, Black L, Blumberg S. Diagnosed Developmental Disabilities in Children Aged 3–17 Years: United States, 2019–2021. National Center for Health Statistics (U.S.); 2023. doi:10.15620/cdc:129520

3. Aldharman SS, Al-Jabr KH, Alharbi YS, et al. Implications of early diagnosis and intervention in the management of neurodevelopmental delay (NDD) in children: A systematic review and meta-analysis. Cureus. 2023;15(5):e38745.

4. Kodesh A, Levine SZ, Khachadourian V, et al. Maternal health around pregnancy and autism risk: a diagnosis-wide, population-based study. Psychol Med. Published online March 26, 2021:1–9.

5. El Marroun H, White TJH, van der Knaap NJF, et al. Prenatal exposure to selective serotonin reuptake inhibitors and social responsiveness symptoms of autism: population-based study of young children. Br J Psychiatry. 2014;205(2):95–102.

6. Moher D, Liberati A, Tetzlaff J, Altman DG, PRISMA Group. Preferred reporting items for systematic reviews and meta-analyses: the PRISMA statement. Int J Surg. 2010;8(5):336–341.

7. Viechtbauer W. Conducting meta-analyses in R with the metafor. J Stat Softw. 2010;36:1–48.

8. Companion R package for the guide Doing Meta-Analysis in R. Accessed March 8, 2024. http://dmetar.protectlab.org/.

9. Shi L, Lin L. The trim-and-fill method for publication bias: practical guidelines and recommendations based on a large database of meta-analyses. Medicine (Baltimore). 2019;98(23):e15987.

10. Wells GA, Shea B, Connell O. The Newcastle-Ottawa Scale (NOS) for Assessing the Quality If Nonrandomized Studies in Meta-Analyses.; 2011.

11. Aromataris E, Munn Z, eds. JBI Manual for Evidence Synthesis. JBI; 2020.

12. Barker TH, Stone JC, Sears K, et al. The revised JBI critical appraisal tool for the assessment of risk of bias for randomized controlled trials. JBI Evid Synth. 2023;21(3):494–506.

13. Nidey NL, Momany AM, Strathearn L, et al. Association between perinatal depression and risk of attention deficit hyperactivity disorder among children: a retrospective cohort study. Ann Epidemiol. 2021;63:1–6.

14. Seebeck J, Sznajder KK, Kjerulff KH. The association between prenatal psychosocial factors and autism spectrum disorder in offspring at 3 years: a prospective cohort study. Soc Psychiatry Psychiatr Epidemiol. Published online August 9, 2023. doi:10.1007/s00127-023-02538-5

15. Higgins JPT. Measuring inconsistency in meta-analyses. BMJ. 2003;327(7414):557-560.

16. Shuffrey LC, Morales S, Jacobson MH, et al. Association of Gestational Diabetes Mellitus and Perinatal Maternal Depression with Early Childhood Behavioral Problems: An Environmental Influences on Child Health Outcomes (ECHO) Study. Child Development. 2023;94(6):1595–1609.

17. Chen LC, Chen MH, Hsu JW, et al. Association of parental depression with offspring attention deficit hyperactivity disorder and autism spectrum disorder: A nationwide birth cohort study. J Affect Disord. 2020;277:109–114.

18. Eilertsen EM, Hannigan LJ, McAdams TA, et al. Parental Prenatal Symptoms of Depression and Offspring Symptoms of ADHD: A Genetically Informed Intergenerational Study. J Atten Disord. 2021;25(11):1554–1563.

19. Olstad EW, Nordeng HME, Sandve GK, Lyle R, Gervin K. Effects of prenatal exposure to (es)citalopram and maternal depression during pregnancy on DNA methylation and child neurodevelopment. Transl Psychiatry. 2023;13(1):149.

20. Van den Bergh BRH, Marcoen A. High antenatal maternal anxiety is related to ADHD symptoms, externalizing problems, and anxiety in 8-and 9-year-olds. Child Dev. 2004;75(4):1085–1097.

21. O’Donnell KJ, Glover V, Lahti J, et al. Maternal prenatal anxiety and child COMT genotype predict working memory and symptoms of ADHD. PLoS One. 2017;12(6):e0177506.

22. Wolford E, Lahti M, Tuovinen S, et al. Maternal depressive symptoms during and after pregnancy are associated with attention-deficit/hyperactivity disorder symptoms in their 3-to 6-year-old children. PLOS ONE. 2017;12(12):e0190248. doi:10.1371/journal.pone.0190248

23. Park M. Maternal Depression Trajectories from Pregnancy to 3 Years Postpartum Are Associated with Children’s Behavior and Executive Functions at 3 and 6 Years. Archives of Women’s Mental Health. Vol 21. #Pages#; 2018.

24. Lahti M, Savolainen K, Tuovinen S, et al. Maternal Depressive Symptoms During and After Pregnancy and Psychiatric Problems in Children. J Am Acad Child Adolesc Psychiatry. 2017;56(1):30–39.e7.

25. Huhdanpää H, Et Aronen P. Prenatal and Postnatal Predictive Factors for Children’s Inattentive and Hyperactive Symptoms at 5 Years of Age: The Role of Early Family-Related Factors. Child Psychiatry and Human Development. Vol 52. #Pages#; 2021.

26. Kingston D, Kehler H, Austin MP, et al. Trajectories of maternal depressive symptoms during pregnancy and the first 12 months postpartum and child externalizing and internalizing behavior at three years. PLoS One. 2018;13(4):e0195365.

27. Koutra K, Roumeliotaki T, Kyriklaki A, et al. Maternal depression and personality traits in association with child neuropsychological and behavioral development in preschool years: Mother-child cohort (Rhea Study) in Crete, Greece. J Affect Disord. 2017;217:89–98.

28. MacKinnon N, Kingsbury M, Mahedy L, Evans J, Colman I. The Association Between Prenatal Stress and Externalizing Symptoms in Childhood: Evidence From the Avon Longitudinal Study of Parents and Children. Biol Psychiatry. 2018;83(2):100–108.

29. Teyhan A, Galobardes B, Henderson J. Child allergic symptoms and mental well-being: the role of maternal anxiety and depression. J Pediatr. 2014;165(3):592–9.e5.

30. Bendiksen B, Aase H, Diep LM, Svensson E, Friis S, Zeiner P. The Associations Between Pre-and Postnatal Maternal Symptoms of Distress and Preschooler’s Symptoms of ADHD, Oppositional Defiant Disorder, Conduct Disorder, and Anxiety. J Atten Disord. 2020;24(7):1057–1069.

31. Van Batenburg-Eddes T, Brion MJ, Henrichs J, et al. Parental depressive and anxiety symptoms during pregnancy and attention problems in children: a crossCcohort consistency study. J Child Psychol Psychiatry. 2013;54(5):591–600.

32. Betts KS, Williams GM, Najman JM, Alati R. Maternal depressive, anxious, and stress symptoms during pregnancy predict internalizing problems in adolescence. Depress Anxiety. 2014;31(1):9–18.

33. Betts KS, Williams GM, Najman JM, Alati R. The relationship between maternal depressive, anxious, and stress symptoms during pregnancy and adult offspring behavioral and emotional problems. Depress Anxiety. 2015;32(2):82–90.

34. Connor O, Ciesla TG, Sefair AA. Maternal prenatal infection and anxiety predict neurodevelopmental outcomes in middle childhood. J Psychopathol Clin Sci. 2022;131:422–434.

35. Breen MS, Wingo AP, Koen N, et al. Gene expression in cord blood links genetic risk for neurodevelopmental disorders with maternal psychological distress and adverse childhood outcomes. Brain Behav Immun. 2018;73:320–330.

36. Yu T, Chang KC, Kuo PL. Paternal and maternal psychiatric disorders associated with offspring autism spectrum disorders: A case-control study. J Psychiatr Res. 2022;151:469–475.

37. Chien YL, Wu CS, Chang YC, Cheong ML, Yao TC, Tsai HJ. Associations between parental psychiatric disorders and autism spectrum disorder in the offspring-A response. Autism Res. 2023;16(5):877–878.

38. Harrington RA, Lee LC, Crum RM, Zimmerman AW. Hertz-Picciotto I. Prenatal SSRI use and offspring with autism spectrum disorder or developmental delay. Pediatrics. 2014;133:e1241–1248.

39. Siracusano M, Riccioni A, Gialloreti LE, et al. Maternal Perinatal Depression and risk of neurodevelopmental disorders in offspring: Preliminary results from the SOS MOOD project. Children (Basel). 2021;8(12):1150.

40. Sørensen MJ, Grønborg TK, Christensen J, et al. Antidepressant exposure in pregnancy and risk of autism spectrum disorders. Clin Epidemiol. 2013;5:449–459.

41. Brown HK, Ray JG, Wilton AS, Lunsky Y, Gomes T, Vigod SN. Association Between Serotonergic Antidepressant Use During Pregnancy and Autism Spectrum Disorder in Children. JAMA. 2017;317(15):1544–1552.

42. Zhou XH, Li YJ, Ou JJ, Li YM. Association between maternal antidepressant use during pregnancy and autism spectrum disorder: an updated meta-analysis. Mol Autism. 2018;9(1). doi:10.1186/s13229-018-0207-7

43. Ramchandani PG, O’Connor TG, Evans J, Heron J, Murray L, Stein A. The effects of pre-and postnatal depression in fathers: a natural experiment comparing the effects of exposure to depression on offspring. J Child Psychol Psychiatry. 2008;49(10):1069–1078.

44. Loomans EM, van der Stelt O, van Eijsden M, Gemke RJBJ, Vrijkotte T, Van den Bergh BRH. Antenatal maternal anxiety is associated with problem behaviour at age five. Early Hum Dev. 2011;87(8):565–570.

45. Chen LM, Pokhvisneva I, Lahti-Pulkkinen M, et al. Independent Prediction of Child Psychiatric Symptoms by Maternal Mental Health and Child Polygenic Risk Scores. Journal of the American Academy of Child & Adolescent Psychiatry. Published online November 2023:S0890856723021858.

46. Le Bas G, Youssef G, Macdonald JA, et al. The role of antenatal and postnatal maternal bonding in infant development. J Am Acad Child Adolesc Psychiatry. 2022;61(6):820–829.e1.

47. Feldman R, Gordon I, Influs M, Gutbir T, Ebstein RP. Parental oxytocin and early caregiving jointly shape children’s oxytocin response and social reciprocity. Neuropsychopharmacology. 2013;38(7):1154–1162.

48. Putnick DL, Sundaram R, Bell EM, et al. Trajectories of maternal postpartum depressive symptoms. Pediatrics. 2020;146(5):e20200857.

49. Tucker JRD, Hobson CW. A systematic review of longitudinal studies investigating the association between early life maternal depression and offspring ADHD. J Atten Disord. 2022;26(9):1167–1186.

50. Maselko J, Sikander S, Bhalotra S, et al. Effect of an early perinatal depression intervention on long-term child development outcomes: follow-up of the Thinking Healthy Programme randomised controlled trial. Lancet Psychiatry. 2015;2(7):609–617.

51. Say GN, Karabekiroğlu K, Babadağı Z, Yüce M. Maternal stress and perinatal features in autism and attention deficit/hyperactivity disorder. Pediatr Int. 2016;58(4):265–269.

52. Gao L, Xi QQ, Wu J, et al. Association between prenatal environmental factors and child autism: A case control study in Tianjin, China. Biomed Environ Sci. 2015;28(9):642–650.

53. Lupattelli A, Chambers CD, Bandoli G, Handal M, Skurtveit S, Nordeng H. Association of maternal use of benzodiazepines and Z-hypnotics during pregnancy with motor and communication skills and attention-deficit/hyperactivity disorder symptoms in preschoolers. JAMA Netw Open. 2019;2(4):e191435.

54. Wiggins LD, Rubenstein E, Daniels J, et al. A phenotype of childhood autism is associated with preexisting maternal anxiety and depression. J Abnorm Child Psychol. 2019;47(4):731–740.

55. Avalos LA, Chandran A, Churchill ML, et al. Prenatal depression and risk of child autism-related traits among participants in the Environmental influences on Child Health Outcomes program. Autism Res. 2023;16(9):1825–1835.

56. Wolf JB, Wade MJ. What are maternal effects (and what are they not)? Philos Trans R Soc Lond B Biol Sci. 2009;364(1520):1107-1115.

57. Odonnell KJ, Glover V, Lahti J. Maternal prenatal anxiety and child COMT genotype predict working memory and symptoms of ADHD. PLoS One. 2017;12(6).

58. Nilsen RM, Vollset SE, Gjessing HK, et al. Self-selection and bias in a large prospective pregnancy cohort in Norway. Paediatr Perinat Epidemiol. 2009;23(6):597–608.

59. Grzeskowiak LE, Morrison JL, Henriksen TB, et al. Prenatal antidepressant exposure and child behavioural outcomes at 7 years of age: a study within the Danish National Birth Cohort. BJOG. 2016;123(12):1919–1928.

60. Lupattelli A, Mahic M, Handal M, Ystrom E, Reichborn-Kjennerud T, Nordeng H. Attention-deficit/hyperactivity disorder in children following prenatal exposure to antidepressants: Results from the Norwegian mother, father, and child cohort study. Obstet Anesth Dig. 2022;42(3):123–123.

61. Bolea-Alamañac B, Davies SJC, Evans J, et al. Does maternal somatic anxiety in pregnancy predispose children to hyperactivity? Eur Child Adolesc Psychiatry. 2019;28(11):1475–1486.

62. D’Souza S, Waldie KE, Peterson ER, Underwood L, Morton SMB. Antenatal and postnatal determinants of behavioural difficulties in early childhood: Evidence from growing up in New Zealand. Child Psychiatry Hum Dev. 2019;50(1):45–60.

63. Huhdanpää H, Morales-Muñoz I, Aronen ET, et al. Prenatal and postnatal predictive factors for children’s inattentive and hyperactive symptoms at 5 years of age: The role of early family-related factors. Child Psychiatry Hum Dev. 2021;52(5):783–799.

64. Hagberg KW, Robijn AL, Jick SS. Maternal depression and antidepressant use during pregnancy and the risk of autism spectrum disorder in offspring. Clin Epidemiol. 2018;10:1599–1612.

65. Clements CC, Castro VM, Blumenthal SR, et al. Prenatal antidepressant exposure is associated with risk for attention-deficit hyperactivity disorder but not autism spectrum disorder in a large health system. Mol Psychiatry. 2015;20(6):727–734.

66. Rai D, Lee BK, Dalman C, Golding J, Lewis G, Magnusson C. Parental depression, maternal antidepressant use during pregnancy, and risk of autism spectrum disorders: population based case-control study. BMJ. 2013;346:f2059.

67. Leis JA, Heron J, Stuart EA, Mendelson T. Associations between maternal mental health and child emotional and behavioral problems: does prenatal mental health matter? J Abnorm Child Psychol. 2014;42(1):161–171.

68. Nishigori T, Hashimoto K, Mori M, et al. Association between maternal prenatal psychological distress and autism spectrum disorder among 3-year-old children: The Japan Environment and Children’s Study. J Dev Orig Health Dis. 2023;14(1):70–76.

69. Hope H, Pierce M, Gabr H, et al. The causal association between maternal depression, anxiety, and infection in pregnancy and neurodevelopmental disorders among 410 461 children: a population study using quasi-negative control cohorts and sibling analysis. Psychol Med. Published online January 11, 2024:1–9.

