## Supplement for "Association of Parental Prenatal Mental Health on Offspring Neurodevelopmental Disorders: A Systematic Review and Meta-Analysis"

### S1. PRISMA checklist

#### *S1.1 PRISMA 2020 for abstracts checklist*

| Section and Topic | Item # | Checklist item | Reported (Yes/No) |
|---|---|---|---|
| **TITLE** | | | |
| Title | 1 | Identify the report as a systematic review. | Yes |
| **BACKGROUND** | | | |
| Objectives | 2 | Provide an explicit statement of the main objective(s) or question(s) the review addresses. | Yes |
| **METHODS** | | | |
| Eligibility criteria | 3 | Specify the inclusion and exclusion criteria for the review. | Yes |
| Information sources | 4 | Specify the information sources (e.g. databases, registers) used to identify studies and the date when each was last searched. | Yes |
| Risk of bias | 5 | Specify the methods used to assess risk of bias in the included studies. | Yes |
| Synthesis of results | 6 | Specify the methods used to present and synthesise results. | Yes |
| **RESULTS** | | | |
| Included studies | 7 | Give the total number of included studies and participants and summarise relevant characteristics of studies. | Yes |
| Synthesis of results | 8 | Present results for main outcomes, preferably indicating the number of included studies and participants for each. If meta-analysis was done, report the summary estimate and confidence/credible interval. If comparing groups, indicate the direction of the effect (i.e. which group is favoured). | Yes |
| **DISCUSSION** | | | |
| Limitations of evidence | 9 | Provide a brief summary of the limitations of the evidence included in the review (e.g. study risk of bias, inconsistency and imprecision). | Yes |
| Interpretation | 10 | Provide a general interpretation of the results and important implications. | Yes |
| **OTHER** | | | |
| Funding | 11 | Specify the primary source of funding for the review. | Yes |
| Registration | 12 | Provide the register name and registration number. | Yes |

*From:* Page MJ, McKenzie JE, Bossuyt PM, Boutron I, Hoffmann TC, Mulrow CD, et al. The PRISMA 2020 statement: an updated guideline for reporting systematic reviews. BMJ 2021;372:n71. doi: 10.1136/bmj.n71

*S.1.2 PRISMA 2020 checklist*

| Section and Topic | Item # | Checklist item | Location where item is reported |
|---|---|---|---|
| **TITLE** | | | |
| Title | 1 | Identify the report as a systematic review. | Title; page 1 |
| **ABSTRACT** | | | |
| Abstract | 2 | See the PRISMA 2020 for Abstracts checklist. | Abstract: page 3; PRISMA checklist: Supplement S1.1 |
| **INTRODUCTION** | | | |
| Rationale | 3 | Describe the rationale for the review in the context of existing knowledge. | Page 5, 6 |
| Objectives | 4 | Provide an explicit statement of the objective(s) or question(s) the review addresses. | Page 5, 6 |
| **METHODS** | | | |
| Eligibility criteria | 5 | Specify the inclusion and exclusion criteria for the review and how studies were grouped for the syntheses. | Page 3 |
| Information sources | 6 | Specify all databases, registers, websites, organisations, reference lists and other sources searched or consulted to identify studies. Specify the date when each source was last searched or consulted. | Page 3 |
| Search strategy | 7 | Present the full search strategies for all databases, registers and websites, including any filters and limits used. | Supplement S2 |
| Selection process | 8 | Specify the methods used to decide whether a study met the inclusion criteria of the review, including how many reviewers screened each record and each report retrieved, whether they worked independently, and if applicable, details of automation tools used in the process. | Page 6, 7 |
| Data collection process | 9 | Specify the methods used to collect data from reports, including how many reviewers collected data from each report, whether they worked independently, any processes for obtaining or confirming data from study investigators, and if applicable, details of automation tools used in the process. | Page 7 |
| Data items | 10a | List and define all outcomes for which data were sought. Specify whether all results that were compatible with each outcome domain in each study were sought (e.g. for all measures, time points, analyses), and if not, the methods used to decide which results to collect. | Table S3 |
| | 10b | List and define all other variables for which data were sought (e.g. participant and intervention characteristics, funding sources). Describe any assumptions made about any missing or unclear information. | Page 6 |

| Section and Topic | Item # | Checklist item | Location where item is reported |
|---|---|---|---|
| Study risk of bias assessment | 11 | Specify the methods used to assess risk of bias in the included studies, including details of the tool(s) used, how many reviewers assessed each study and whether they worked independently, and if applicable, details of automation tools used in the process. | Page 7, 8 |
| Effect measures | 12 | Specify for each outcome the effect measure(s) (e.g. risk ratio, mean difference) used in the synthesis or presentation of results. | Page 7 |
| Synthesis methods | 13a | Describe the processes used to decide which studies were eligible for each synthesis (e.g. tabulating the study intervention characteristics and comparing against the planned groups for each synthesis (item #5)). | Table S1 |
| | 13b | Describe any methods required to prepare the data for presentation or synthesis, such as handling of missing summary statistics, or data conversions. | Page 7, 8 |
| | 13c | Describe any methods used to tabulate or visually display results of individual studies and syntheses. | Page 7, 8 |
| | 13d | Describe any methods used to synthesize results and provide a rationale for the choice(s). If meta-analysis was performed, describe the model(s), method(s) to identify the presence and extent of statistical heterogeneity, and software package(s) used. | Page 7, 8 |
| | 13e | Describe any methods used to explore possible causes of heterogeneity among study results (e.g. subgroup analysis, meta-regression). | Page 7, 8 |
| | 13f | Describe any sensitivity analyses conducted to assess robustness of the synthesized results. | Page 8 |
| Reporting bias assessment | 14 | Describe any methods used to assess risk of bias due to missing results in a synthesis (arising from reporting biases). | Page 14 |
| Certainty assessment | 15 | Describe any methods used to assess certainty (or confidence) in the body of evidence for an outcome. | Table S2 |
| **RESULTS** | | | |
| Study selection | 16a | Describe the results of the search and selection process, from the number of records identified in the search to the number of studies included in the review, ideally using a flow diagram. | Page 8 |
| | 16b | Cite studies that might appear to meet the inclusion criteria, but which were excluded, and explain why they were excluded. | None |
| Study characteristics | 17 | Cite each included study and present its characteristics. | Page 8-14; Table S1-S3 |
| Risk of bias in studies | 18 | Present assessments of risk of bias for each included study. | Figure S2-S3 |
| Results of individual studies | 19 | For all outcomes, present, for each study: (a) summary statistics for each group (where appropriate) and (b) an effect estimate and its precision (e.g. confidence/credible interval), ideally using structured tables or plots. | Figure 2 and 3 |

| Section and Topic | Item # | Checklist item | Location where item is reported |
|---|---|---|---|
| Results of syntheses | 20a | For each synthesis, briefly summarise the characteristics and risk of bias among contributing studies. | Page 14 |
| | 20b | Present results of all statistical syntheses conducted. If meta-analysis was done, present for each the summary estimate and its precision (e.g. confidence/credible interval) and measures of statistical heterogeneity. If comparing groups, describe the direction of the effect. | Figure 2 and 3 |
| | 20c | Present results of all investigations of possible causes of heterogeneity among study results. | Page 9, 10, 11 |
| | 20d | Present results of all sensitivity analyses conducted to assess the robustness of the synthesized results. | Page 9, 10, 11 |
| Reporting biases | 21 | Present assessments of risk of bias due to missing results (arising from reporting biases) for each synthesis assessed. | Figures S3, S5 |
| Certainty of evidence | 22 | Present assessments of certainty (or confidence) in the body of evidence for each outcome assessed. | |
| **DISCUSSION** | | | |
| Discussion | 23a | Provide a general interpretation of the results in the context of other evidence. | Page 15 |
| | 23b | Discuss any limitations of the evidence included in the review. | Page 15-16 |
| | 23c | Discuss any limitations of the review processes used. | |
| | 23d | Discuss implications of the results for practice, policy, and future research. | Page 16 |
| **OTHER INFORMATION** | | | |
| Registration and protocol | 24a | Provide registration information for the review, including register name and registration number, or state that the review was not registered. | Page 6 |
| | 24b | Indicate where the review protocol can be accessed, or state that a protocol was not prepared. | Page 6 |
| | 24c | Describe and explain any amendments to information provided at registration or in the protocol. | Supplement S3 |
| Support | 25 | Describe sources of financial or non-financial support for the review, and the role of the funders or sponsors in the review. | Pages 19 |
| Competing interests | 26 | Declare any competing interests of review authors. | Page 19 |
| Availability of data, code and other materials | 27 | Report which of the following are publicly available and where they can be found: template data collection forms; data extracted from included studies; data used for all analyses; analytic code; any other materials used in the review. | |

*From:* Page MJ, McKenzie JE, Bossuyt PM, Boutron I, Hoffmann TC, Mulrow CD, et al. The PRISMA 2020 statement: an updated guideline for reporting systematic reviews. BMJ 2021;372:n71. doi: 10.1136/bmj.n71

### S2. Database search

#### *S2.1 PubMed*

We used the following combination of keywords for the PubMed search:

(Prenatal[title] OR Antenatal[title] OR perinatal[title] OR Maternal[title] OR Paternal[Title] OR Parental[title]) AND (ADHD[title/abstract] OR (attention deficit/hyperactivity disorder[Title/Abstract]) OR (attention deficit hyperactivity[Title/Abstract]) OR (attention deficit disorder[Title/Abstract]) OR ADD[Title/Abstract] OR Hyperactive[Title/Abstract] OR inattention[Title/Abstract] OR inattentive [Title/Abstract] OR ASD[Title/Abstract] OR (Autism spectrum disorder[Title/Abstract]) OR Autism[Title/Abstract] OR autistic[Title/Abstract] OR Aspergers[Title/Abstract] OR atypical[Title/Abstract] OR (intellectual disability[Title/Abstract]) OR (Neurodevelopmental Disorder[Title/Abstract]) OR (Mental Retardation[Title/Abstract])) AND (anxiety[Title/Abstract] OR (generalized anxiety disorder[Title/Abstract]) OR GAD[Title/Abstract] OR anxious[Title/Abstract] OR phobia[Title/Abstract] OR agoraphobia[Title/Abstract] OR (adjustment disorder[Title/Abstract]) OR phobic[Title/Abstract] OR anxiety[Title/Abstract] OR (panic disorder[Title/Abstract]) OR (post-traumatic stress[Title/Abstract]) OR (selective mutism[Title/Abstract]) OR OCD[Title/Abstract] OR (Obsessive-compulsive disorder[Title/Abstract]) OR (Obsessive-compulsive[Title/Abstract]) OR obsession[Title/Abstract] OR obsessive[Title/Abstract] OR Compulsion[Title/Abstract] OR Compulsive[Title/Abstract] OR Depression[Title/Abstract] OR (Major Depression[Title/Abstract]) OR (Major Depressive Disorder[Title/Abstract]) OR MDD[Title/Abstract] OR (Pervasive Depressive Disorder[Title/Abstract]) OR PDD[Title/Abstract] OR Depressed[Title/Abstract] OR Depressive[Title/Abstract]) AND (Kid[Title/Abstract] OR Kids[Title/Abstract] OR offspring[Title/Abstract] OR child[Title/Abstract] OR children[Title/Abstract] OR kin[Title/Abstract] OR son[Title/Abstract] OR daughter[Title/Abstract]) NOT rats[Title] NOT rat[Title] NOT dog[Title] NOT dogs[Title] NOT rabbits[Title] NOT rabbit[Title] NOT mice[Title] NOT mouse[title]

#### *S2.2 CENTRAL*

We used the following combination of keywords for the Cochrane Central Register of Controlled Trials (CENTRAL) search:

(title record) Prenatal OR Antenatal OR perinatal OR Maternal OR Paternal OR Parental
AND
(Title & abstract) ADHD OR "attention deficit hyperactivity disorder" OR "attention deficit hyperactivity" OR "attention deficit disorder" OR ADD OR Hyperactive OR inattention OR inattentive OR ASD OR "Autism spectrum disorder" OR Autism OR autistic OR Aspergers OR atypical OR "intellectual disability" OR "Neurodevelopmental Disorder" OR "Mental Retardation"
AND
(Title & abstract) anxiety OR "generalized anxiety disorder" OR GAD OR anxious OR phobia OR agoraphobia OR "adjustment disorder" OR phobic OR "panic disorder" OR "post-traumatic stress" OR "selective mutism" OR OCD OR "Obsessive-compulsive disorder" OR "Obsessive-compulsive" OR obsession OR obsessive OR Compulsion OR Compulsive OR Depression OR "Major Depression" OR "Major Depressive Disorder" OR MDD OR "Pervasive Depressive Disorder" OR PDD OR Depressed OR Depressive
AND
(Title & abstract) Kid OR offspring OR child OR children OR kin OR son OR daughter
NOT
(Title & abstract) rats AND dog AND rabbits AND mice AND animal

*S2.3 OVID*

We used the following combination of keywords for the OVID search:

Prenatal OR Antenatal OR perinatal OR Maternal Or Paternal OR Parental
AND
ADHD OR attention deficit hyperactivity disorder OR attention deficit hyperactivity OR attention deficit disorder OR Hyperactive OR inattention OR inattentive
OR
ASD OR Autism spectrum disorder OR Autism OR autistic OR Aspergers OR atypical
OR
intellectual disability OR Neurodevelopmental Disorder OR Mental Retardation
AND
anxiety OR generalized anxiety disorder OR GAD OR anxious OR phobia OR agoraphobia OR adjustment disorder OR phobic OR panic disorder OR post-traumatic stress OR selective mutism
OR
OCD OR Obsessive-compulsive disorder OR Obsessive-compulsive OR obsession OR obsessive OR Compulsion OR Compulsive
OR
Depression OR Major Depression OR Major Depressive Disorder OR MDD OR Pervasive Depressive Disorder OR PDD OR Depressed OR Depressive
AND
Kid OR offspring OR child OR children OR kin OR son OR daughter
NOT
rat NOT dog NOT rabbits NOT mice NOT mouse NOT animal

*S2.4 Google Scholar*

We used the following combination of keywords for the Google Scholar search. We screened the first 100 articles to identify any relevant articles that had been missed in previous searches.

(intitle:Prenatal OR intitle:Antenatal OR intitle:perinatal OR intitle:Maternal OR intitle:Paternal OR intitle:Parental) AND (ADHD OR "attention deficit/hyperactivity disorder" OR "attention deficit hyperactivity" OR "attention deficit disorder" OR ADD OR Hyperactive OR inattention OR inattentive OR ASDOR "Autism spectrum disorder" OR Autism OR autistic OR Aspergers OR atypical OR "intellectual disability" OR "Neurodevelopmental Disorder" OR "Mental Retardation")
AND
(anxiety OR "generalized anxiety disorder" OR GAD OR anxious OR phobia[Title/Abstract] OR agoraphobia OR "adjustment disorder" OR phobic OR anxiety OR "panic disorder" OR "post-traumatic stress" OR "selective mutism" OR OCD OR "Obsessive-compulsive disorder" OR "Obsessive-compulsive" OR obsession OR obsessive OR Compulsion OR Compulsive OR Depression OR "Major Depression" OR "Major Depressive Disorder" OR MDD OR "Pervasive Depressive Disorder" OR PDD OR Depressed OR Depressive)
AND
(Kid OR Kids OR offspring OR child OR children OR kin OR son OR daughter)

### S3. Narrative synthesis of studies, methods

In our narrative synthesis, we adopted a structured approach to reporting, consistent with the Synthesis without meta-analysis (SWiM) guidelines.[1]

*Grouping studies for synthesis*

For the narrative synthesis, where available, we included available information on both prenatal and postnatal diagnoses in parents and NDDs to discuss differences in the associations between these disorders. We also examined methodological differences between studies to better understand factors contributing to associations between PMADs and NDDs.

Two researchers (APK and LEC) conducted full-text screenings of the articles selected for the narrative synthesis to extract any additional descriptive statistics and summary information on research findings. The narrative synthesis was initially structured according to the type of parent exposure, as prespecified in the PROSPERO protocol. Following full-text article screenings, one author (APK) identified additional groupings based on how frequently they were discussed in preserved articles. These extra groupings were added to provide additional insights on outcomes of interest (NDDs) and study designs. The groupings were added if they were present in at least two articles to ensure that study groupings were sufficiently large for synthesis. The groupings included: (1) differences in NDD presentation based on offspring sex/gender, given gender/sex differences in frequency and symptoms of NDDs; (2) types of PMAD and NDD measurements, to demonstrate how measurement may contribute to heterogeneity between findings; (3) timing of PMAD measurement, prenatally or prenatally and postnatally, to explore potential differences in associations depending on the timing of PMAD assessment and identify sensitive periods for the impact of PMADs on NDDs; and (4) factors impacting the association between PMADs and NDDs, to summarize what factors are most frequently implicated and the quality of evidence on these factors. These additional groups were jointly agreed upon by the research team. One researcher (APK) organized study findings by groups, and two authors (LEC and BM) jointly reviewed if studies assigned to each group were relevant.

*Synthesis methods*

Data in narrative synthesis were not transformed statistically, owing to a variety of statistical methods and designs in reviewed studies (Supplemental Table 1, column Study design). Our review presents how many studies reported congruent findings in order to facilitate easy identification of relevant studies. However, we did not conduct vote counting based on direction of effect. We have also refrained from conducting sign tests for evaluating differences between proportions of positive and negative directions of effect reported in synthesized studies. [2] This decision aligns with a recent critique that the sign test assumption (that analyzed data have a binomial distribution) may not accurately represent underlying structures of data in narrative syntheses.[3] Instead, our methods of synthesis included tables of descriptives, ordered by parent exposure, offspring outcome, study sample size (Supplemental Table 1), and risk of bias (Supplemental Table 2). We report types of PMAD and NDD measurements in Supplemental Table 1.

*Criteria used to prioritize results for summary and synthesis*

We prioritized studies which addressed our groupings (*Grouping studies for synthesis*), in particular themes of clinical relevance: (1) sex/gender differences in NDD presentation; (2) studies which explicitly addressed how the timing of parental exposure may impact offspring outcomes; (3) mediators and specific factors impacting the association between PMADs and NDDs. We also highlighted studies which discussed methodological issues potentially relevant to quality of multiple studies (e.g., parental rater bias in assessing offspring behaviour). We did not prioritize studies based on risk of bias assessments, owing to the fact that the number of available studies was small for several exposures (parental OCD; paternal PMADs) and outcomes (offspring ID; sex/gender-specific NDD effects in the offspring). In those cases, we included all available studies.

*Investigation of heterogeneity in reported effects*

We have organized study tables by potential sources of heterogeneity: sample sizes (Table S1) and risk of bias (Table S2). We did not order study tables by commonly selected subpopulations of age and sex for two reasons. First, detailed age data for parents and offspring were inconsistently reported across studies. Second, there was a limited number of studies directly comparing effects of exposures between parents or assessing sex/gender differences in offspring outcomes. The investigation of heterogeneity was not pre-specified in the meta-analysis PROSPERO protocol and was specified at the data extraction stage, based on discussion and consensus between co-authors.

*Assessing certainty of evidence*

We used Newcastle-Ottawa scales to assess the quality of evidence in case-control and cohort studies. Our modification of the scales included adding a criterion of whether studies accounted for potential genetic covariates shared by parents and offspring, and whether studies erroneously used fathers as a control for samples of mothers with PMADs. We used the Joanna Briggs Institute (JBI) Critical Appraisal Checklists to assess the quality of cross-sectional studies and a randomized controlled trial. Two authors (APK and BM) independently assessed risk of bias and resolved any disagreements through discussion.

*Data presentation methods*

We summarized studies in the narrative synthesis with study tables (Table S1, Table S2). In Table S1, we included key characteristics: parent exposure; time of parent exposure measurement; parent age; measures of exposure; offspring outcomes; offspring age; measures of offspring outcomes; study designs; sample sizes, including broken down by offspring sex/gender; setting; name of study or register, where applicable; and breakdown of samples by participant race, country of origin, ancestry, or ethnicity, whichever was reported. In Table S2, we divided studies by risk of bias.

### S4. Risk of bias assessment scales

#### *S4.1 Scoring for a modified Newcastle-Ottawa (NOS) Scale for Case-Control Studies*

A star represents an allocated point. Possible score range: 0-9 (two points maximum can be given for control for confounders). Score categorization: ≥8 = high quality study (low risk of bias), 7 to 6 = moderate quality study (moderate risk of bias), ≤5 = low quality study (high risk of bias).

Selection

Adequate case definition:

a) independent validation (>1 person/record/time/process to extract data) *
b) record linkage/self-report
c) no description

Representativeness of the cases:

a) all eligible cases of parents with PMADs and children with NDDs, or sample representative of different ages, ethnicities, genders, etc. or appropriate random sample *
b) selection bias, not representative, or not stated

Selection of controls:

a) controls from the same community, matched by age and sex/gender *
b) controls not matched
c) no description

Definition of controls:

a) controls without psychiatric diagnoses or matched for age and comorbidity status (if participants have multiple occurrences of outcome) *
b) not stated or fathers used as controls for mothers

Comparability of Cohorts

Control for confounders:

a) study adjusts for genetic factors shared between parents and offspring *
b) study controls for any additional factors or provides sensitivity analyses *
c) no description

Exposure

Ascertainment:

a) interviewer blind to case/control status *
b) interviewer not blind to case/control status
c) self-report or medical record only
d) no description

Method of ascertainment:

a) same for cases and controls *
b) different assessment procedures
c) no description

Non-response rate:

a) same rate for cases and controls *
b) non-responders described or groups different
c) not stated

#### *S4.2 Scoring for a modified Newcastle-Ottawa (NOS) Scale for Cohort Studies*

A star represents an allocated point. Possible score range: 0-10 (two points maximum can be given for control for confounders). Score categorization: ≥8 = high quality study (low risk of bias), 7 to 6 = moderate quality study (moderate risk of bias), ≤5 = low quality study (high risk of bias).

Selection

Representativeness of the exposed cohort:

a) truly representative of average individuals with PMAD *
b) somewhat representative of average individuals with PMADs *
c) selected group of individuals with PMADs
d) no description of the derivation of the cohort

Selection of the non-exposed cohort:

a) drawn from the same community as the exposed cohort *
b) drawn from a different source
c) no description of the derivation of the non-exposed cohort

Ascertainment of exposure:

a) secure record *
b) structured interview *
c) written self-report
d) no description

Demonstration that outcome of interest (offspring neurodevelopmental disorder/symptom) was not present at start of study:

a) yes *
b) no

Comparability of cohorts

Control for confounders:

a) study adjusts for genetic factors shared between parents and offspring *
b) study controls  for any additional factors or provides sensitivity analyses *
c) no description

Outcome
Ascertainment:

a) independent blind assessment *
b) record linkage *
c) interviewer not blinded, self-report or medical record only

d) no description

Was follow-up long enough for outcomes to occur:

a) yes *
b) no

Adequacy of follow up of cohorts:

a) complete follow up - all subjects accounted for *
b) subjects lost to follow up unlikely to introduce bias - small number lost to follow up, or description provided of those lost *
c) low follow-up rate or and no description of those lost
d) no statement

#### *S4.3 Modified Joanna Briggs Institute (JBI) Critical Appraisal Checklist for Analytical Cross Sectional Studies*

Each question can be answered yes, no, unclear, or not/applicable.

1. Were the criteria for inclusion in the sample clearly defined?
2. Were the study subjects and the setting described in detail (demographics, location, and time period)?
3. Was the exposure measured in a valid and reliable way (is the measure a gold standard measure or compared to a gold standard and are measurements consistent across repetitions)?
4. Were objective, standard criteria used for measurement of the condition (is participant selection based on specified diagnosis, clear definition, or matching participants by key characteristics)?
5. Were confounding factors identified (were they measured)?
6. Were strategies to deal with confounding factors stated (e.g., matching or stratifying patients or the quality of regression analysis to account for confounders)?
7. Were the outcomes measured in a valid and reliable way (diagnostic criteria, validated instruments, self-report? Were data collectors comparable in levels of education, research experience etc.)?
8. Was appropriate statistical analysis used (how were confounders measured, were strata of data defined by the specified variables, was analytical strategy appropriate in terms of its assumptions)?

#### *S4.4 Joanna Briggs Institute (JBI) Critical Appraisal Tool for Assessment of Risk of Bias for Randomized Controlled Trials 2023*

Each question can be answered yes, no, unclear, or not/applicable.

Question 1: Was true randomization used for assignment of participants to treatment groups?

Question 2: Was allocation to groups concealed?

Question 3: Were treatment groups similar at the baseline?

Question 4: Were participants blind to treatment assignment?

Question 5: Were those delivering the treatment blind to treatment assignment?

Question 6: Were treatment groups treated identically other than the intervention of interest?

Question 7: Were outcome assessors blind to treatment assignment?

Question 8: Were outcomes measured in the same way for treatment groups?

Question 9: Were outcomes measured in a reliable way?

Question 10: Was follow up complete and if not, were differences between groups in terms of their follow up adequately described and analysed?

Question 11: Were participants analysed in the groups to which they were randomized?

Question 12: Was appropriate statistical analysis used?

Question 13: Was the trial design appropriate and any deviations from the standard RCT design (individual randomization, parallel groups) accounted for in the conduct and analysis of the trial?

**Figure S1.** Funnel plot for studies addressing maternal prenatal mood and anxiety disorders and offspring neurodevelopmental disorders.

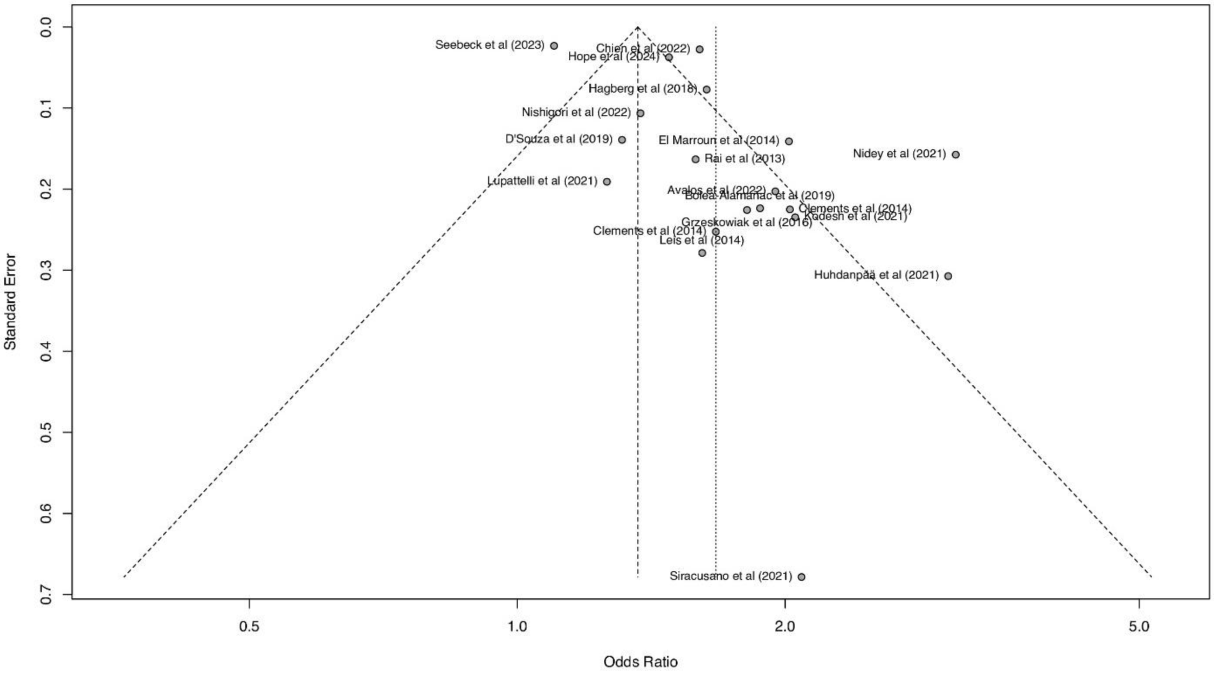

Data points represent original studies in the meta-analysis.

**Figure S2.** Forest plot of associations between maternal prenatal mood and anxiety disorders and offspring neurodevelopmental disorders, with outlier studies removed.

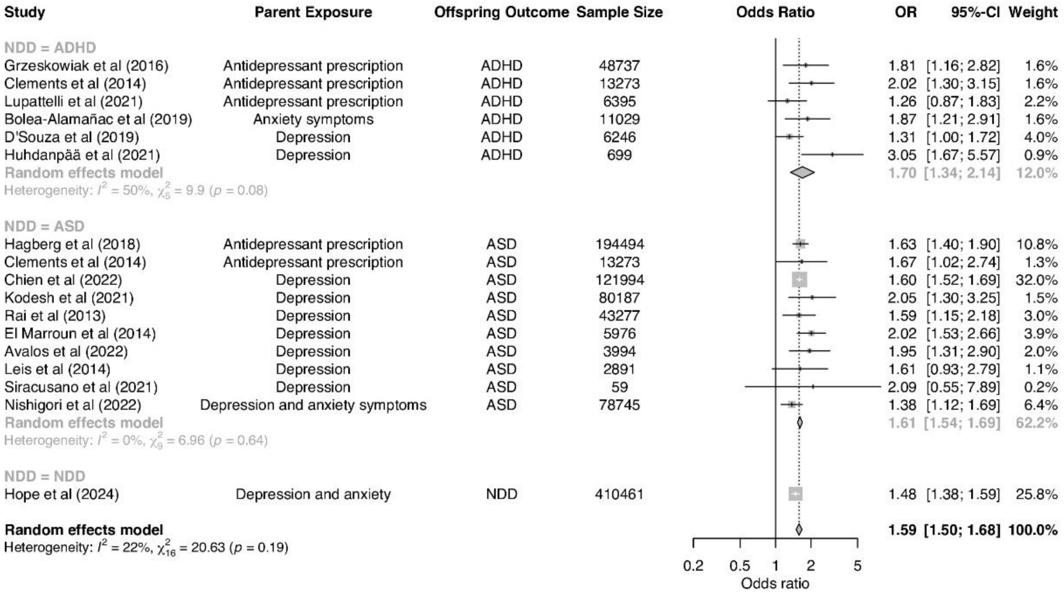

Studies deemed to be outliers were Nidey et al (2021) and Seebeck et al (2023) because their effect sizes differed largely from the overall effect in the meta-analysis.

Square sizes reflect the weights attributed to each study. Diamonds denote the summary effect sizes for the random-effect models.
CI=confidence intervals ADHD=attention-deficit/hyperactivity disorder. ASD=autism spectrum disorder. NDD=neurodevelopmental disorders.
*Clements et al (2015) analyzed separate samples of individuals with diagnoses of autism spectrum disorder and attention-deficit/hyperactivity disorder. Authors provided results for three trimesters. The third trimester has been selected for this meta-analysis because it is the most conservative estimate reported.
†Chien et al (2022) included the following disorders: major depressive disorder, persistent depressive disorder, and depressive disorder, unspecified.
‡Hope et al (2024) analyzed a combined sample of individuals with any of the following disorders: autism/autism spectrum disorder, attention-deficit/hyperactivity disorder, intellectual disability, cerebral palsy, and epilepsy.

**Figure S3.** Contour-enchanced funnel plot using the trim and fill method for studies addressing maternal prenatal mood and anxiety disorders and offspring neurodevelopmental disorders.

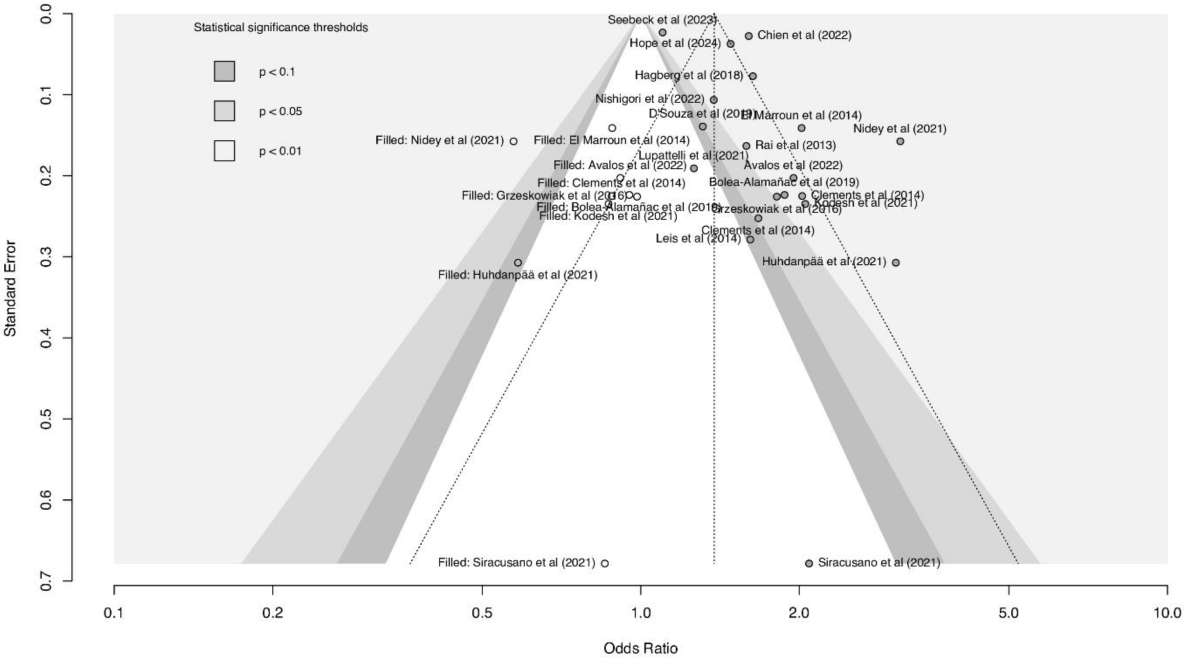

Gray data points represent original studies in the meta-analysis and white data points represent filled studies, i.e., studies simulated by the trim and fill method. Shaded regions represent traditional thresholds below and of statistical significance (p-values) to illustrate whether results of imputed studies would be statistically significant or not.

**Figure S4.** Funnel plot for studies addressing paternal prenatal mood and anxiety disorders and offspring neurodevelopmental disorders.

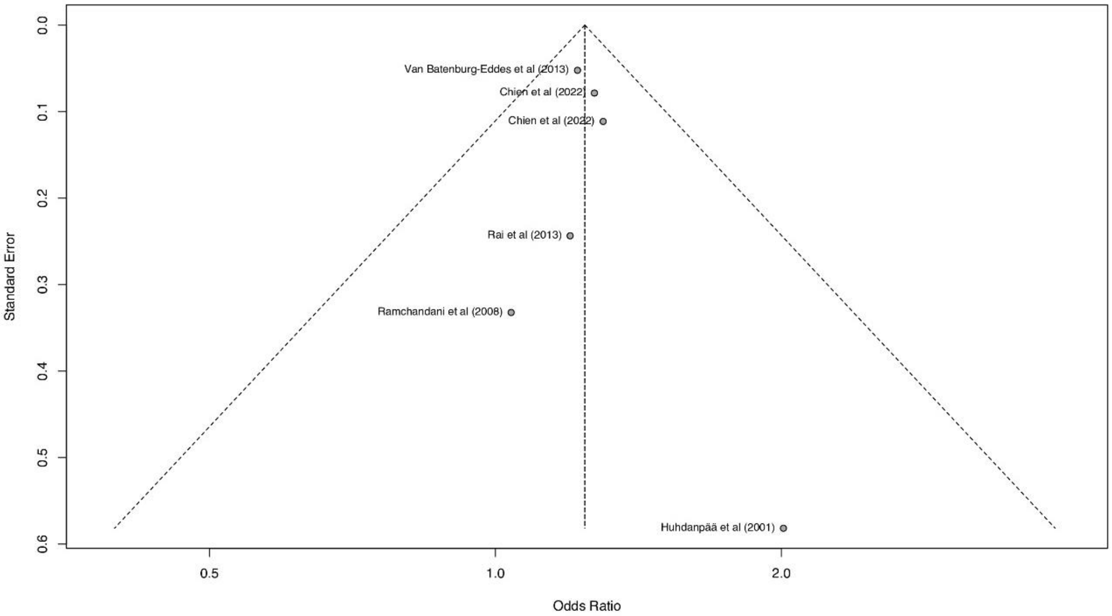

Data points represent original studies in the meta-analysis.

**Figure S5.** Contour-enchanced funnel plot using the trim and fill method for studies addressing paternal prenatal mood and anxiety disorders and offspring neurodevelopmental disorders.

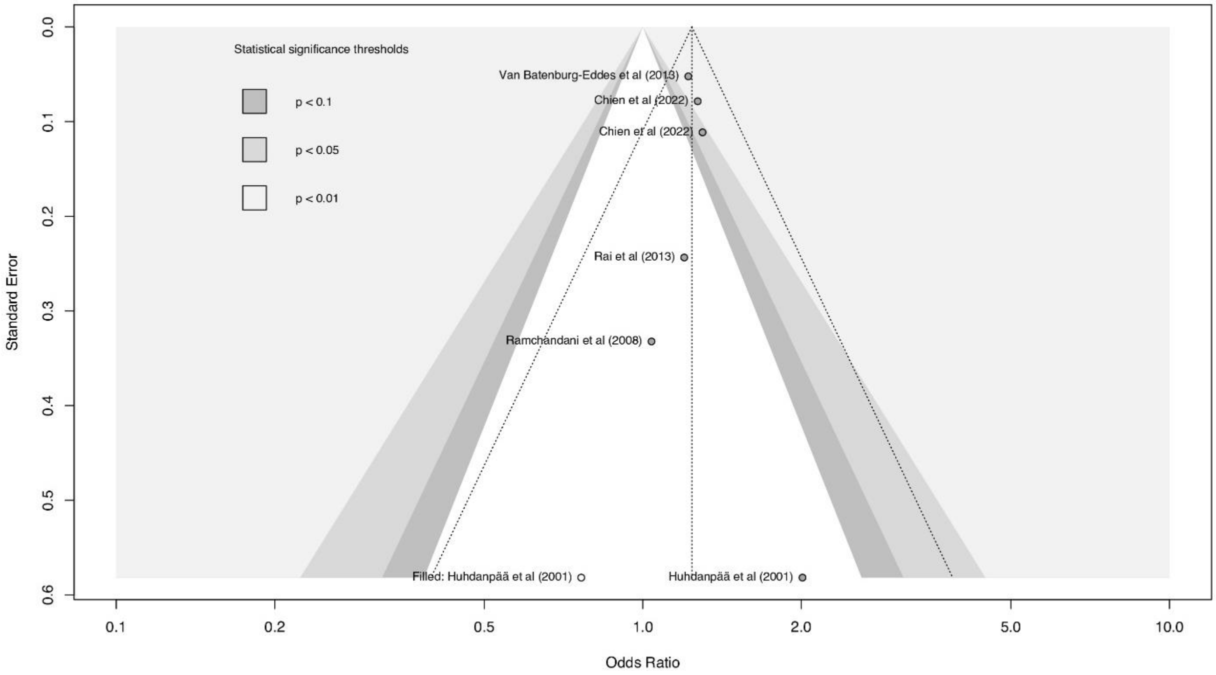

Gray data points represent original studies in the meta-analysis and white data points represent filled studies, i.e., studies simulated by the trim and fill method. Shaded regions represent traditional thresholds below and of statistical significance (p-values) to illustrate whether results of imputed studies would be statistically significant or not.

### References

1 Campbell M, McKenzie JE, Sowden A, *et al*. Synthesis without meta-analysis (SWiM) in systematic reviews: reporting guideline. *BMJ* 2020; **368**: l6890.

2 Chapter 12: Synthesizing and presenting findings using other methods. https://training.cochrane.org/handbook/current/chapter-12 (accessed March 9, 2024).

3 Nikolakopoulos S. Misuse of the sign test in narrative synthesis of evidence. *Res Synth Methods* 2020; **11**: 714–9.
